# JUNIPER: Reconstructing Transmission Events from Next-Generation Sequencing Data at Scale

**DOI:** 10.1101/2025.03.02.25323192

**Authors:** Ivan Specht, Gage K. Moreno, Taylor Brock-Fisher, Lydia A. Krasilnikova, Brittany A. Petros, Jonathan E. Pekar, Mark Schifferli, Ben Fry, Catherine M. Brown, Lawrence C. Madoff, Meagan Burns, Stephen F. Schaffner, Daniel J. Park, Bronwyn L. MacInnis, Al Ozonoff, Patrick Varilly, Michael D. Mitzenmacher, Pardis C. Sabeti

## Abstract

Transmission reconstruction––the inference of who infects whom in disease outbreaks––offers critical insights into how pathogens spread and provides opportunities for targeted control measures. We developed JUNIPER (Joint Underlying Network Inference for Phylogenetic and Epidemiological Reconstructions), a highly-scalable pathogen outbreak reconstruction tool that incorporates intrahost variation, incomplete sampling, and algorithmic parallelization. Central to JUNIPER is a statistical model for within-host variant frequencies observed by next generation sequencing, which we validated on a dataset of over 160,000 deep-sequenced SARS-CoV-2 genomes. Combining this within-host variation model with population-level evolutionary and transmission models, we developed a method for inferring phylogenies and transmission trees simultaneously. We benchmarked JUNIPER on computer-generated and real outbreaks in which transmission links were known or epidemiologically confirmed. We demonstrated JUNIPER’s real-world utility on two large-scale datasets: over 1,500 bovine H5N1 cases and over 13,000 human COVID-19 cases. Based on these analyses, we quantified the elevated H5N1 transmission rates in California and identified high-confidence transmission events, and demonstrated the efficacy of vaccination for reducing SARS-CoV-2 transmission. By overcoming computational and methodological limitations in existing outbreak reconstruction tools, JUNIPER provides a robust framework for studying pathogen spread at scale.

## INTRODUCTION

Advances in pathogen genomic surveillance have enhanced our ability to determine infectious disease transmission pathways from the accumulation of mutations over time [1–20].

Reconstructed transmission networks provide critical insight into how pathogens spread, quantify key parameters such as the evolution rate and effective reproductive number, and permit the evaluation of mitigation strategies such as vaccination [16] or nonpharmaceutical interventions [21,22].

Numerous algorithms for reconstructing outbreaks based on epidemiological and consensus-level genomic data have been proposed [1,4,9,12,14,19,20,23], but few account for the additional evidence for transmission provided by within-host variants. Broadly, consensus-based tools quantify the likelihood of transmission for each pair of cass based on the single nucleotide variant (SNV) distance and the difference in time of sampling. While a lower SNV distance at the consensus level is a stronger indicator of transmission than a higher one, consensus-based approaches do not model intrahost single nucleotide variants (iSNVs), which are known to be key indicators of transmission links [10,17,24]. Specifically, the presence of a certain genome sequence as a minor variant in one case and as the consensus genome in another makes it more likely that these cases form a donor-recipient pair––one in which the donor transmitted the minor variant to the recipient. Some transmission inference methods incorporate iSNVs into phylogenetic inference at both the intrahost and population levels [10,25], but they require lengthy compute time that is intractable for large datasets, e.g. over 100 cases.

Existing methods also face limitations in capturing the statistical properties of sampling mechanisms and transmission patterns. Some methods assume that all cases in an outbreak are detected and sequenced, which is rare for datasets from large outbreaks or for pathogens that frequently infect asymptomatically [11,26]. Other methods infer the number of unsampled hosts along the transmission chain from one sequenced case to another, but do not model the full underlying transmission tree––a simplification that introduces biases in parameter estimates.

Moreover, most methods are not adequately designed to quantify the impact of superspreading as they do not explicitly model overdispersion in transmission networks, i.e., a small number of cases are responsible for a large proportion of transmissions [27].

With these current limitations in mind, we conceptualized JUNIPER as a unifying framework for extracting transmission links from any genomic-epidemiological dataset. We first developed a statistical model for the frequencies of iSNVs in next-generation sequencing (NGS) data, enabling JUNIPER to infer transmissions more reliably. We then approached the problem of missing data by letting JUNIPER infer both the known and unknown positive cases in an outbreak, as well as the probability of sampling each case. To enable JUNIPER to scale to large datasets, we implemented a parallelization method and a customized Markov Chain Monte Carlo (MCMC) algorithm. For validation, we generated synthetic datasets and sought out real datasets with known or strongly-suspected transmission links, giving us a ground truth against which to compare JUNIPER’s inferences. Finally, using JUNIPER, we examined geographic regions of elevated H5N1 avian flu transmission amid the ongoing cattle in the United States outbreak and quantified the effect of COVID-19 vaccination on transmission.

**Figure 1:**
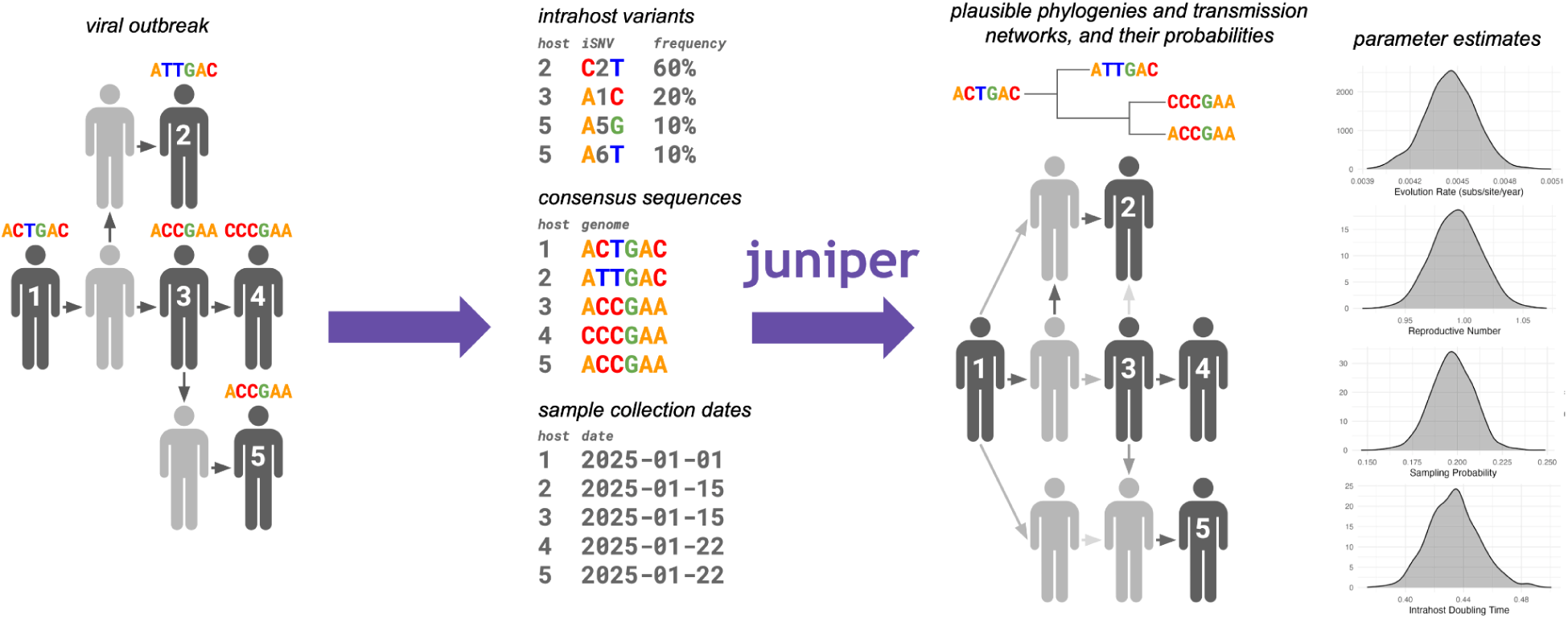
Overview of JUNIPER. JUNIPER takes in consensus genomes, sample collection dates, and (optionally) within-host variants from fully- or partially-sampled outbreaks. It conducts Bayesian inference on the space of possible transmission networks and underlying phylogenetic trees, as well as genomic and epidemiological parameters such as the evolution rate and reproductive number.

## RESULTS

### Modeling Within-Host Variants

We first developed a statistical model to describe the proportion of the intrahost viral population exhibiting a given iSNV. In brief, we model the within-host effective population size after inoculation as a pure-birth process, a population model where the birth rate of virions far exceeds the death rate. In this model, the effective population size within a host *t* days after inoculation is calculated as exp(β*t*), with β being the growth rate parameter in units of days^-1^. Once the first mutation event occurs at a given site on the viral genome, the proportion *x* of virions exhibiting a *de novo* iSNV is inversely related to the population size when the mutation occurred, assuming there are no selective pressures and that sites evolve independently. Letting μ represent the mutation rate in substitutions per site per day and setting *r* = μ/β, the resulting probability density function of *x* is

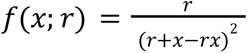

(see **Methods**). We note that when a minor allele frequency threshold is applied, such as *x* > 3% as is common in NGS data to filter out erroneous iSNV calls, the renormalized density function is approximately proportional to 1/*x*^2^ for small *r*.

To validate our statistical model for *de novo* iSNV frequencies, we visualized all iSNV frequencies observed across a dataset of 166,521 SARS-CoV-2 genomes from Massachusetts, USA, mainly of the Delta and Omicron variants (see **Methods**). We observed minimal discrepancy between the empirical probability density and the theoretical probability density function, *f*(*x*) ∝ 1/*x*^2^ over the support *x* > 3% (**Figure 2**). In addition to reliably capturing the data distribution, our model can also make inferences about within-host evolutionary parameters. We estimated *r* to be 2.70 × 10^-6^ by maximum likelihood. While the parameter *r* itself does not have a clear biological interpretation, it helps us quantify the evolution rate and/or the intrahost population growth rate because it equals the ratio of these two quantities. Existing studies estimate the evolution rate μ to be on the order of 10^-6^ substitutions per site per day [28–31], with higher estimates variants of concern [32]. With μ = 4.2 × 10^-6^ (95% HPD: [1.7 × 10^-6^, 2.0 × 10^-5^]) substitutions per site per day as estimated for the Delta variant [33], we obtain a β of 1.6 (95% HPD: [0.63, 7.4]). Values of β within this HPD are consistent with observed viral loads, estimated to be 10^6^–10^9^ virions [28] at peak infectiousness, which on average occurs approximately 7 days post-inoculation [34].

**Figure 2:**
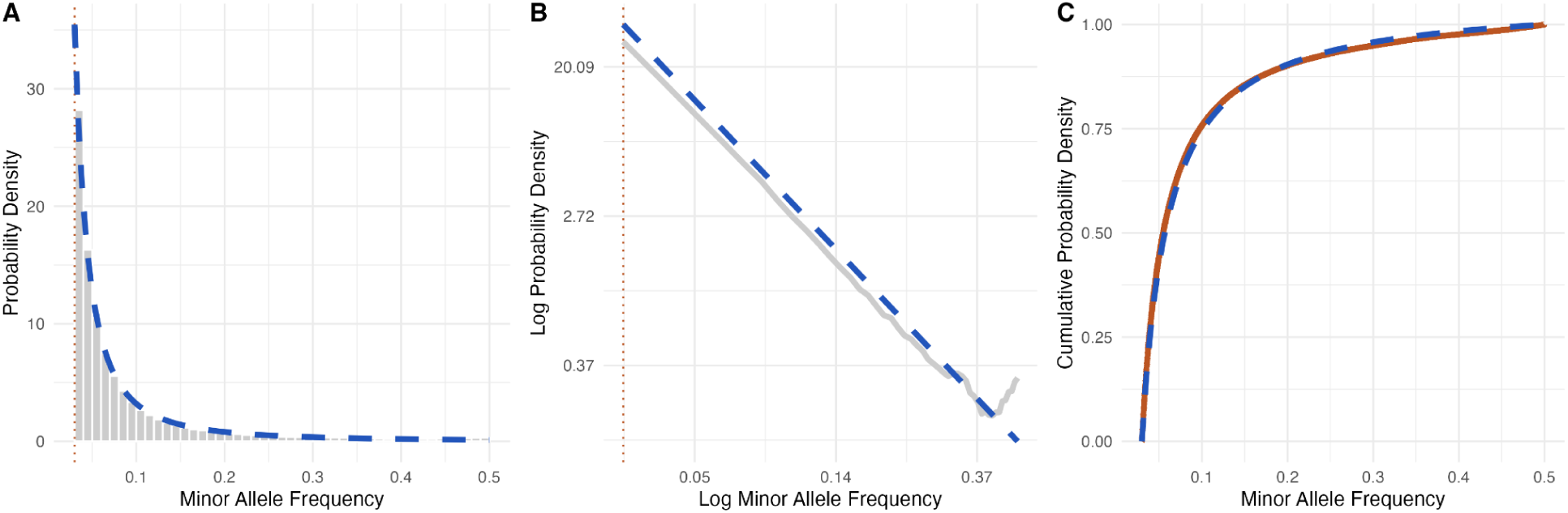
Modeling minor allele frequencies. **(A)** Histogram of minor allele frequencies from the dataset of 166,521 genomes (gray bars) and modeled probability density function (blue dashed line). **(B)** Empirical (gray) and theoretical probability density functions. **(C)** Empirical cumulative density function (orange solid line) and modeled cumulative density function (blue dashed line). A minor allele frequency filter of at least 3% was applied for both figures.

### Reconstructing Phylogenetic and Transmission Trees

By combining our model for iSNVs frequencies with existing models of molecular evolution and epidemic spread, we developed a joint likelihood function for the consensus genomes, within-host variants, and sample collection times. We modeled transmission as a random branching process originating from one infectious individual, where the number of individuals infected by each positive case, the generation intervals (difference in times of infection for a donor-recipient pair), and sojourn intervals (difference in time of sampling and infection) are drawn from prespecified probability distributions. To calculate the likelihood associated with all donor-recipient pairs, infection times, and sampling times in an outbreak, we adapted the method of TransPhylo [9], an outbreak reconstruction tool that accounts for both sampled and unsampled cases. This method assigns a probability to any proposed transmission tree linking the sampled cases by integrating over all possible trees that contain the proposed one as a subtree, invoking numerical methods to do so.

We designed JUNIPER to infer both transmission networks and phylogenetic trees simultaneously, which requires a phylogenetic tree likelihood function, in addition to an epidemiological one. This is in contrast to TransPhylo, which is limited by the fact that the user must prespecify a fixed phylogenetic tree. To relate transmission trees to phylogenetic trees, we assumed that each host is inoculated by a single viral particle, which is tenable for a range of viruses known to have tight bottlenecks. We further treated all coalescences on the underlying phylogeny as occurring approximately at the time of inoculation of a host, which follows from our assumption that the within-host effective population size grows rapidly following inoculation. These assumptions completely determine the shape of the tree, conditional on the transmission network, so that we need not vary the network and tree separately. We quantified the phylogenetic tree likelihood using the Jukes-Cantor model [35]. JUNIPER can optionally infer incomplete bottlenecks, but we note this setting currently requires making several additional assumptions and approximations that may result in overconfidence in the existence of transmission links between hosts with shared intra-host variants. (See <u>Supplementary Text,</u> <u>Section A.4</u> for derivation.)

To handle large datasets, we engineered JUNIPER to run in parallel across CPU cores. JUNIPER does so by partitioning the proposed transmission network into subtrees, performing MCMC on each subtree on a different core simultaneously, then reassembling the trees and repeating with a different partition. This approach is made possible by the fact that in JUNIPER, the posterior probability calculated for each subtree in a partition does not depend on that of any other subtree. By contrast, standard Bayesian phylogenetic tree reconstruction requires computing the coalescent prior, which does not have the property that its value on one subtree is unaffected by its value on a different subtree. JUNIPER replaces the population and sampling constraints captured by the coalescent prior with the aforementioned transmission tree likelihood, which factors over subtrees and hence gives rise to seamless parallelization.

The JUNIPER codebase also makes use of efficient data structures and MCMC moves. Unlike nearly all existing methods, JUNIPER stores the exact timing of each mutation along each branch of the underlying phylogeny as an unknown parameter. This idea, first proposed by Nielsen et al. [36], allows for fast computation of the phylogenetic tree likelihood––a speedup that outweighs the increased dimensionality of the parameter space. Finally, JUNIPER employs numerous custom MCMC moves, including ones that propose substantial rearrangements of the transmission network in order to reduce the total number of mutations on the underlying phylogenetic tree. These moves, detailed in <u>Supplementary Text, Section B</u>, accelerate JUNIPER’s ability to find high-likelihood configurations of the parameter space.

### Validation on Simulated Data

To measure JUNIPER’s performance and compare it to existing methods, we simulated 23 synthetic outbreaks with varying evolutionary parameters, transmission dynamics, and sampling probabilities (see **Supplementary Table S1** and ***Synthetic Data Generation*** in **Methods**). We designed these datasets to reflect real-world scenarios, in which a significant fraction of cases may go undetected and genome sequences may contain missing nucleotide calls. We assessed the reconstruction tools with three metrics: the average probability assigned to the true donor of each recipient (*donor-recipient accuracy*), the fraction of recipients whose true donor is contained in the smallest set of hosts with an inferred >95% chance of containing the donor (*donor set accuracy*), and the fraction of recipients for which the most probable inferred donor is indeed the true donor (*most likely donor accuracy*). We computed these statistics based on direct transmissions as well as indirect ones, meaning transmissions among observed hosts with unsampled intermediates in between. Note that for direct transmission links, we treated all unsampled intermediates as indistinguishable, meaning that if the true donor of a case is unobserved, then the contribution to each accuracy metric is calculated based on the probability of that case having *any* unobserved ancestor. We quantified the accuracy of the tools’ parameter estimates, e.g. the inferred evolution rates, using root mean square error (RMSE).

We reconstructed the synthetic outbreaks using JUNIPER under four settings (targeted inputs, default inputs, consensus genomes only, and without inferring unsampled intermediates), and in comparison to three current methods: TransPhylo [37] (with the underlying phylogenetic tree generated via IQ-TREE [38,39] and passed through TreeTime [40]), outbreaker2 [41], and BadTrIP [10] (which, unlike outbreaker2 or TransPhylo, incorporates iSNVs). By *targeted inputs*, we mean that the user-provided generation interval distribution and sojourn interval distribution matched the distributions under which the data were generated. This would represent situations in which we know the pathogen and its epidemiological characteristics with high confidence. We also assessed JUNIPER’s robustness to model misspecification by running the model with *default inputs*, in which we set the generation and sojourn interval to Gamma distributions each with mean and variance of five days, regardless of the data-generating distributions. To quantify the impact of within-host variation data for inferring transmission links in JUNIPER, we ran it with consensus genomes alone for comparison. Lastly, we ran JUNIPER without the ability to infer any unsampled intermediates in order to assess the importance of modeling missing data.

Across all experiments, JUNIPER consistently outperformed the other methods in terms of both transmission link reconstruction and parameter estimation (see **Figure 3** and **Supplementary Figure S1**). This held true for both targeted and default JUNIPER inputs, with only a marginal decrease in accuracy for the latter. On average across experiments, the donor-recipient accuracy for JUNIPER with targeted inputs was highest, at 52.2% for direct transmission links and 51.0% for indirect transmission links (see **Figure 3A–B**). Our synthetic datasets often contained multiple positive cases with near-identical genomes and similar sampling times, making the exact infector of each recipient challenging to determine from within a small set of plausible infectors and hence resulting in modest donor-recipient accuracies. Donor set accuracy, which measures the reliability of these sets, was once again the highest for JUNIPER with targeted inputs at 99.2% for direct transmissions and 95.6% for indirect transmissions (see **Figure 3A–B**). Although specific donor-recipient pairs may be challenging to extract from sets of plausible ones, we found that JUNIPER with targeted inputs most often assigns the highest probability over possible donors to the correct donor for each recipient (see **Figure 3A–B**). This metric, captured through most likely donor accuracy, equalled 63.9% for direct transmissions and 60.9% for indirect transmissions. Across all three metrics, JUNIPER with default inputs was only about 1% less accurate as compared to targeted inputs. All other methods exhibited significantly lower accuracy.

**Figure 3:**
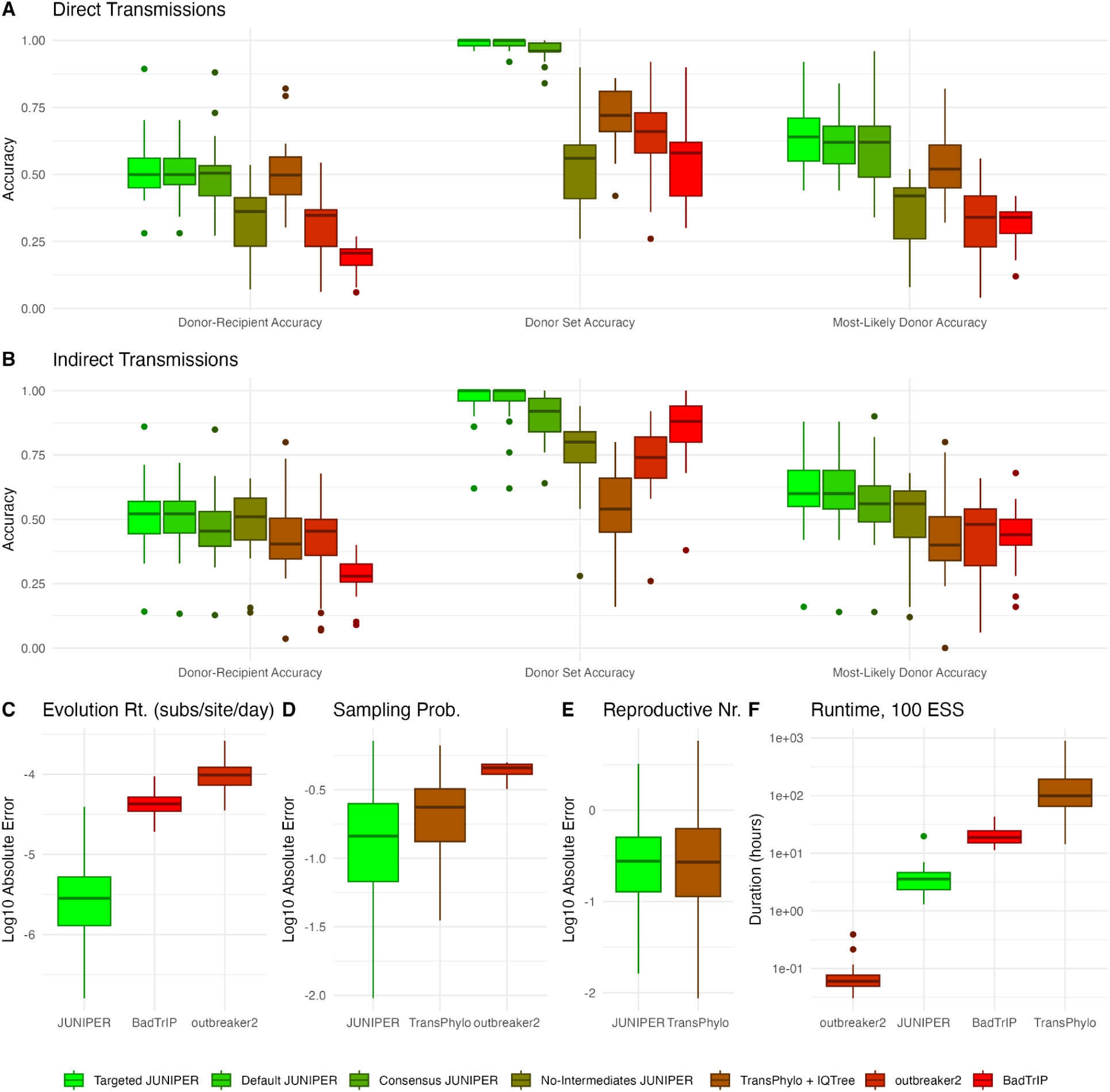
Assessing JUNIPER’s performance on simulated data, aggregated across experiments. **(A–B)** Donor-recipient accuracy, donor set accuracy, and most-likely donor accuracy computed based on **(A)** direct transmission links and **(B)** direct and indirect transmissions links. Results are compared for JUNIPER with targeted input parameters (light green), JUNIPER with default input parameters (medium green), JUNIPER without access to within-host variation data (dark green), JUNIPER under the assumption of no unsampled intermediates (green-brown), TransPhylo and IQ-TREE (brown), outbreaker2 (crimson), and BadTrIP (red). **(C–E)** Base-10 log absolute error (inferred parameter values in posterior samples minus true parameter values) in estimating the **(C)** mutation rate, **(D)** sampling rate, and **(E)** reproductive number for applicable methods. The reproductive number and epidemic start date are not inferred by outbreaker2; the evolution rate and epidemic start date are not inferred by IQ-TREE or TransPhylo; and only the mutation rate is inferred by BadTrIP. **(F)** Runtime to achieve an effective sample size of 100 for JUNIPER with targeted input parameters, TransPhylo and IQ-TREE, outbreaker2, and BadTrIP. All statistics for BadTrIP were computed based only on 21 of 23 synthetic outbreaks, as the presence of any ambiguous sites resulted in prohibitively long runtimes, estimated at 23 days per 100 effective samples for the remaining two.

JUNIPER consistently exhibited the least error in estimating the mutation rate, sampling rate, and reproductive number, often by a wide margin (see **Figure 3C–E**). JUNIPER was the only method that inferred all of these values, and we compare our results to other methods as applicable. Averaged over experiments and over posterior samples, the RMSE for JUNIPER with targeted inputs estimating the mutation rate was 6.00 × 10^-6^ subs/site/day, with all simulated mutation rates being on the order of 10^-5^ subs/site/day. For comparison, the RMSE for BadTrIP and outbreaker2 were 668% and 1,800% greater than for JUNIPER, respectively. This severe error may be attributed in part to the fact that outbreaker2 can only model coalescences occurring in sampled hosts, and that BadTrIP did not infer the existence of any unsampled intermediates across all experiments despite having the ability to do so. These limitations resulted in BadTrIP and outbreaker2 inferring less total evolutionary time and hence an elevated mutation rate to compensate. TransPhylo does not infer mutation rates. JUNIPER estimated the sampling probability with a RMSE of 0.209. The RMSE for this parameter was 27.3% higher for TransPhylo and 117% higher for outbreaker2, again attributable to the limited space of possible transmission networks outbreaker2 is able to sample. The RMSE in estimating the reproductive number was 0.499 for JUNIPER; for TransPhylo it was 92.6% greater. The runtime for JUNIPER averaged 2.36 hours per 100 effective samples, significantly faster than the BadTrIP’s average of 21.3 hours per 100 effective samples and TransPhylo’s average of 194 hours per 100 effective samples based on the log posterior (see **Supplementary Figure 3F**). JUNIPER ran slower than outbreaker2, whose runtime averaged 0.0792 hours per 100 effective samples, respectively (see **Figure 3F**). This slowdown is attributable in part to JUNIPER’s use of iSNV data and ability to infer any transmission network topology with sampled and unsampled cases, in contrast to outbreaker2.

We further identified several scenarios in which the accuracy of JUNIPER remained consistent, but the accuracy or performance of other methods significantly decreased. Outbreaker2 performed markedly worse in the presence of any sequencing error, whereas other methods exhibited only a marginal decrease in accuracy (see **Supplementary Figure S1Q**, experiments 22–23). The runtime for BadTrIP was prohibitively long in the presence of any ambiguous sites on the genome, estimated at 23 days per 100 effective samples. Therefore, we only assessed BadTrIP’s performance and computed summary statistics based on experiments 1–21. We note that for all experiments, the direct or indirect transmissions that may be inferred by TransPhylo are restricted compared to the other methods, since TransPhylo overlays transmission events on top of a fixed, user-provided phylogenetic tree. This prohibits links incompatible with the user-provided tree while boosting the probabilities of compatible ones; as a result, we saw more extreme probabilities (approximately 0 or 1) assigned to true transmission links in TransPhylo than in other methods (see **Supplementary Figure S1P**).

### Validation on Real Data

We sought out real-world genomic data in the form of controlled experiments or densely-sampled outbreaks with known transmission routes to validate JUNIPER further and benchmark it against other methods. While few such datasets exist, we identified a controlled study of bovine respiratory syncytial virus (RSV) in Swedish cows [18] and a rigorously-investigated SARS-CoV-2 outbreak at a hospital in South Africa [42,43] as strong candidates for methodological validation on real data.

We reconstructed both outbreaks using JUNIPER, outbreaker2, BadTrIP, and TransPhylo (with an underlying phylogeny generated by IQ-TREE and TreeTime). We examined JUNIPER under three settings: *standard settings*, *incomplete bottlenecks*, and *no intermediates*. ‘Standard settings’ is the analog of targeted inputs from the *in silico* validation, in which we selected the epidemiological parameters (e.g. generation interval) to match those of the pathogen as closely as possible. We also ran JUNIPER using the optional feature of inferring ‘incomplete bottlenecks’ (see <u>Supplementary Text, Section A.4</u>), which is useful for datasets with extremely low diversity at the consensus level. In the bovine RSV dataset, all consensus sequences were identical, so shared minor variants and temporal information were the only basis for transmission inference. Finally, we ran JUNIPER under the assumption that ‘no intermediates’ were unsampled. In contrast to the synthetic datasets, all cases were sampled and sequenced in the bovine RSV dataset, as were nearly all known cases in the SARS-CoV-2 dataset. Prohibiting JUNIPER from inferring undetected cases may lead to higher accuracy in such scenarios, and permits fair comparison to tools that do not infer unsampled intermediates.

We first assessed the methods in the bovine RSV controlled study, where true transmission links were known. A total of 9 cows were infected through co-housing or by obtaining bronchoalveolar lavages from infectious animals, with each successive transmission event occurring seven days after the previous one. All cows were otherwise housed in separate pens to avoid unintended transmission events [18] (see **Figure 4A–B**). JUNIPER with the incomplete bottleneck setting performed best in terms of donor-recipient accuracy (i.e. average probability that an inferred transmission link is true) at 59.5%. JUNIPER under all three settings had accuracies higher than other methods, at 55.1%, 53.8%, 44.5%, 40.1%, and 25.4%, for standard JUNIPER, JUNIPER with no intermediates, outbreaker2, BadTrIP, and TransPhylo, respectively.

**Figure 4:**
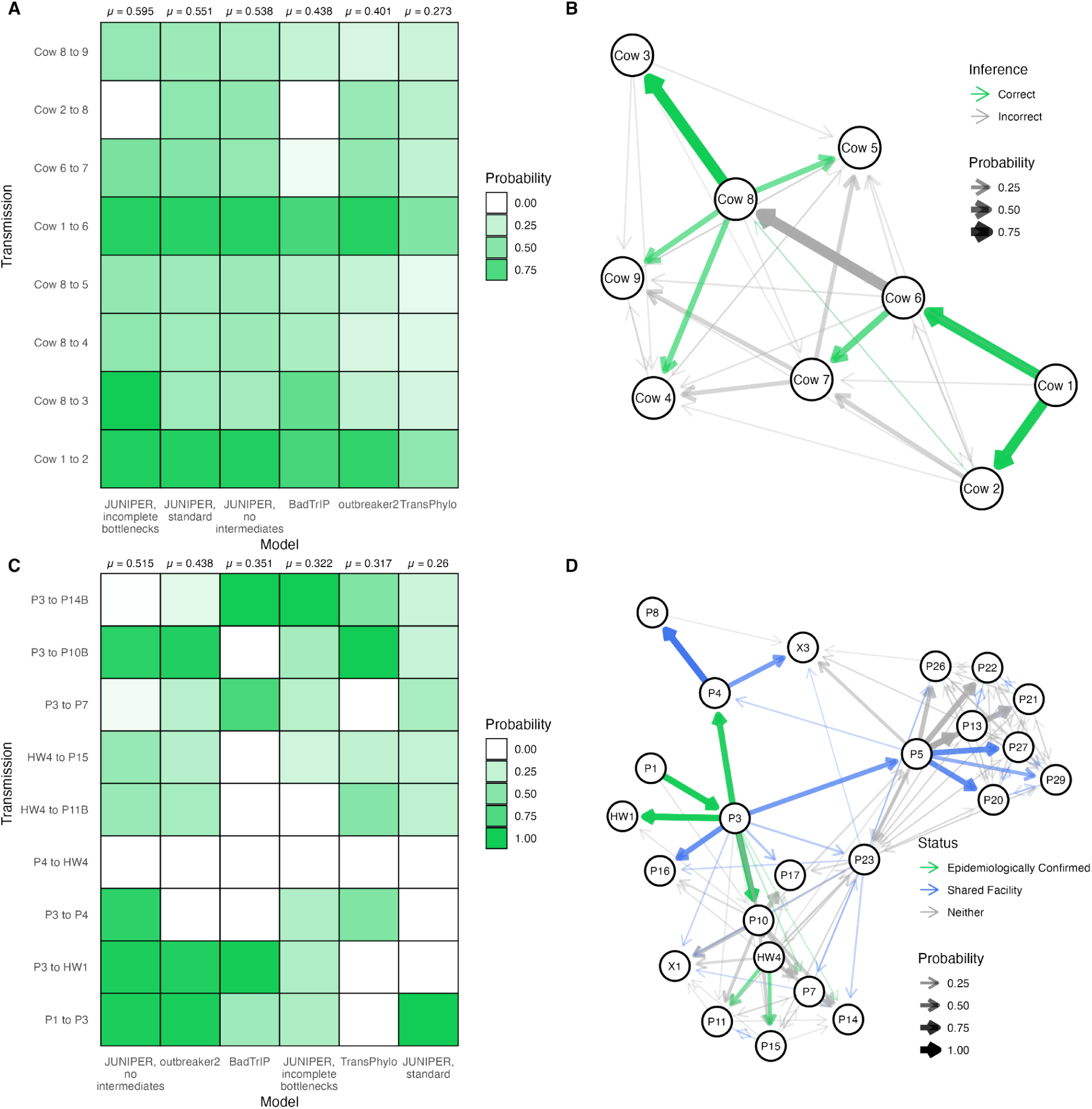
Assessing JUNIPER’s performance on a controlled bovine RSV outbreak and an epidemiologically-investigated SARS-CoV-2 outbreak. **(A)** Posterior probabilities assigned to each of the true transmission links in the bovine RSV using JUNIPER (under three different settings), outbreaker2, BadTrIP, and TransPhylo. **(B)** Inferred transmission links for the bovine RSV outbreak according to JUNIPER with the incomplete bottleneck inference setting, with opacity corresponding to posterior probability. True transmission links shown in green. **(C)** Posterior probabilities assigned to each of the transmission links in the South Africa SARS-CoV-2 outbreak deemed likely by epidemiological investigation using JUNIPER (under three different settings), outbreaker2, BadTrIP, and TransPhylo. **(D)** Inferred transmission links for the SARS-CoV-2 outbreak according to JUNIPER under the assumption of no unsampled intermediates, with opacity corresponding to posterior probability. Likely transmission links according to epidemiological investigation shown in green (see **Supplementary Table S2**); transmissions between patients residing in the same hospital facility shown in blue.

We next applied the same methods to the nosocomial SARS-CoV-2 outbreak in South Africa [42,43]. Among the 39 infected persons, 9 transmission links between patients with sequenced samples were deemed highly probable by epidemiological investigation, giving us a benchmark against which to assess all methods (see **Supplementary Table S2**). In addition to epidemiologically-confirmed links, patient movement histories throughout the hospital were documented, suggesting certain transmissions to be plausible based on patient co-location. Here, instructing JUNIPER to assume all cases were sequenced performed best overall based on the epidemiologically-confirmed links, while the accuracies of other JUNIPER methods underperformed existing tools. The average probability that an inferred transmission link was an an epidemiologically-identified transmission link was 51.5%, 43.8%, 35.1%, 32.2%, 31.7%, and 26.0% for JUNIPER with complete sampling, outbreaker2, BadTrIP, JUNIPER with incomplete bottleneck inference, TransPhylo, and JUNIPER with standard settings, respectively (see **Figure 4C–D**). These probabilities serve as the analog of donor-recipient accuracy when true links are unknown but epidemiologically-likely ones have been proposed, allowing us to assess how well the methods match manual outbreak investigation. Manual and genomic epidemiological analyses may disagree; for instance, all six methods refute the P4 to HW4 link (see **Figure 4C–D**), but it was reported that the two patients came into close contact several times when P4 was coughing (see **Supplementary Table S2**).

### Application to Large-Scale H5N1 and SARS-CoV-2 Datasets

Outbreak reconstruction at the population scale––made possible by JUNIPER’s ability to run MCMC in parallel––can provide critical insights about pathogen spread when combined with additional data, including location and vaccination status. We obtained thousands of genomes from the ongoing avian influenza outbreak in United States cattle and the COVID-19 pandemic to identify geographical regions with elevated transmission rates and to measure the effect of vaccination on the number of transmissions per positive case, respectively. Such findings help public health authorities best allocate resources to high-transmission settings and advocate for the adoption of pharmaceutical interventions.

We first used JUNIPER to analyze 1,519 H5N1 genomes collected between March and December 2024 (see **Figure 5A–C**) [44]. By computing the expected number of transmissions per sampled case by state (**Figure 5B**), we found the highest reproductive number in California, estimated at 1.28 (95% HPD: 1.26–1.29) as compared to the overall inferred reproductive number of 1.06 (95% HPD: 1.03–1.09). This is consistent with a sharp increase in California samples we observed in December 2024, and reflects the state of emergency due to H5N1 declared by California’s government on December 18th, 2024. Among the California samples, JUNIPER identified 32 putative transmission links with posterior probability exceeding 50%, suggesting targets of further case investigation (**Figure 5C**). We were able to achieve effective sample sizes exceeding 50 for each inferred parameter over a total runtime of approximately 3 days and 5 hours using a 32-CPU core Linux virtual machine, highlighting JUNIPER’s ability to conduct near real-time transmission inference at scale. For additional posterior parameter densities, see **Supplementary Figure S2**; for an assessment of MCMC convergence, see **Supplementary Figure S3**.

**Figure 5:**
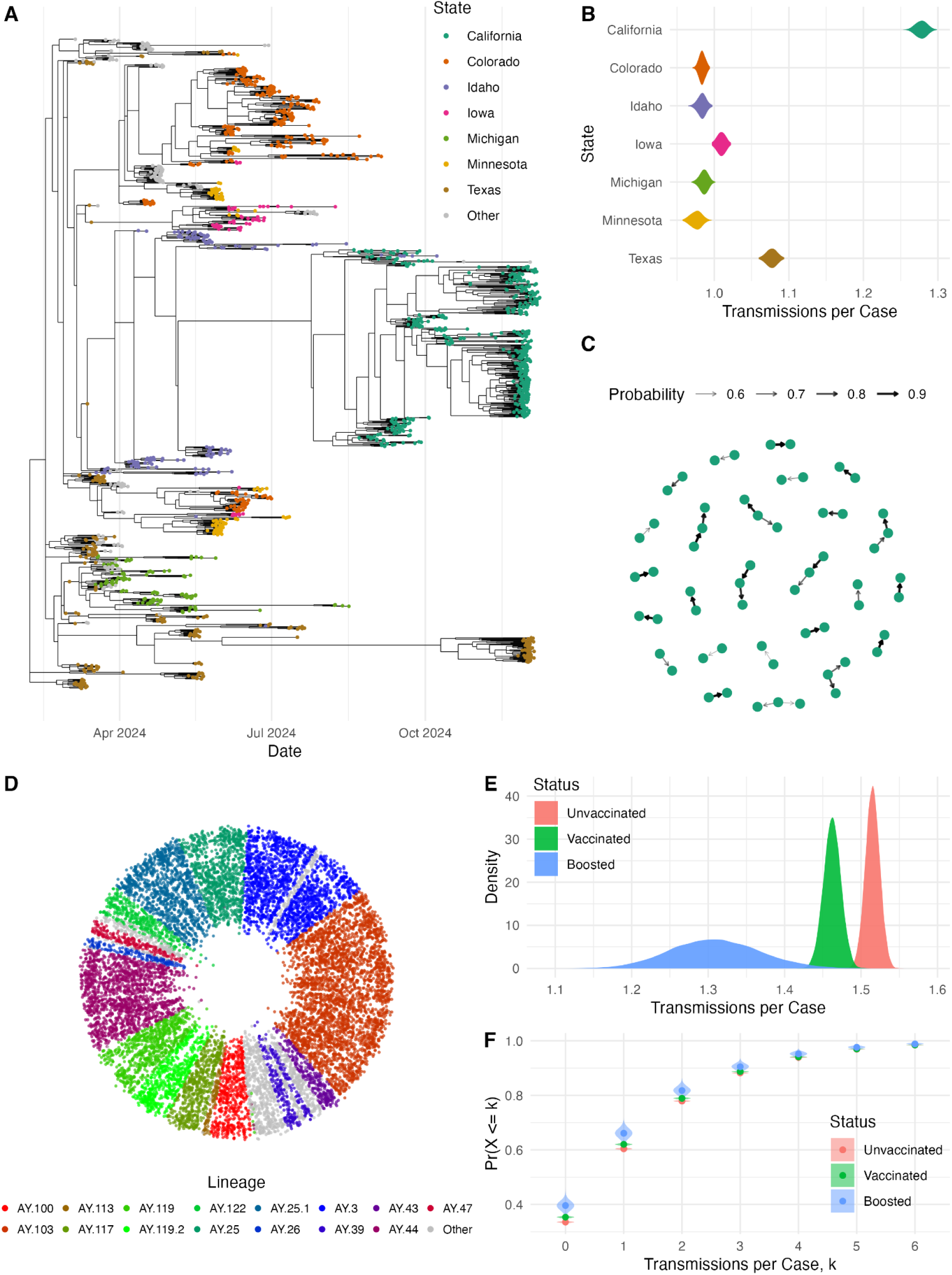
Analyzing H5N1 and SARS-CoV-2 Transmission with JUNIPER. **(A)** A single posterior transmission network sample for 1,519 H5N1 cases detected in U.S. cattle between March and December 2024. Nodes represent sampled cases, and edges represent direct or indirect transmission links. Unsampled hosts are not shown. Geographic location is shown for each sampled case. **(B)** Posterior densities of the expected number of transmissions per case by state. **(C)** Transmission network plot for the California samples. Only direct transmissions between sampled cattle with posterior probability exceeding 50% are shown. **(D)** A single posterior transmission network sample for 13,570 SARS-CoV-2 genomes from Massachusetts, USA, collected in November 2021. Cases that are inferred to have transmitted to one another are positioned adjacent to each other, and the inferred time of infection is represented as the distance from the center of the circle. Nodes are colored by lineage with unsampled hosts not shown. **(E)** Posterior densities of expected transmissions per person based on vaccination status. **(F)** Posterior densities of the empirical cumulative distribution function of the number of transmissions per person based on vaccination status.

Finally, we conducted large-scale transmission reconstruction with JUNIPER to examine the effect of COVID-19 vaccination on the number of infectees per infector. Ground-truth transmission networks of SARS-CoV-2 are practically impossible to determine with certainty at scale, so JUNIPER provides the only probabilistic approach to this investigation. We analyzed 13,570 SARS-CoV-2 genomes from Massachusetts, USA, in November 2021, when Delta variant case counts were growing exponentially (see **Figure 5D–F**). For each case, various metadata were provided by the Massachusetts Department of Public Health, including vaccination status. By superimposing these data on posterior transmission network samples, we found a highly significant difference in transmission rate based on vaccination status. Boosted individuals transmitted at a 34.1% lower rate than unvaccinated individuals, and vaccinated (but not boosted) individuals transmitted at a 9.81% lower rate than unvaccinated individuals.

Although we cannot conclude causality from these results due to possible confounding effects (e.g. vaccinated people having fewer social contacts than non-vaccinated people), the differences in transmission rate were highly statistically significant as shown by **Figure 5E**. The total runtime was approximately 6 days and 17 hours using a Linux virtual machine with 224 CPU cores, achieving an effective sample size of 82 based on the log posterior and over 100 for each inferred parameter (see **Supplementary Figure S4**). For further analysis of the underlying epidemic parameters such as the evolution rate, see **Supplementary Figure S2**.

## DISCUSSION

JUNIPER advances the state of the art in transmission reconstruction by incorporating within-host variants, modeling missing data, and scaling to large, sparsely sampled datasets. We validated and benchmarked JUNIPER on synthetic, controlled, and epidemiologically-investigated case studies, demonstrating robust performance across diverse scenarios. JUNIPER exhibited a marked improvement over existing methods in terms of both its accuracy and its applicability to large and sparse datasets. JUNIPER’s analysis of the 1,519 H5N1 genomes quantified differences in transmission patterns and risk by geographic location, allowing for tailored public health response. Our JUNIPER run on 13,570 Massachusetts SARS-CoV-2 genomes illustrated how transmission inference at the population level provides crucial insights into the efficacy of interventions such as vaccination. Such near real-time analyses can empower public health authorities to make data-driven decisions to curtail future spread.

JUNIPER uses both consensus-level genomes and within-host variants to predict transmission links, resulting in more accurate inferences than existing methods. This feature is made possible by our novel statistical model of *de novo* iSNV frequencies, which we combine with existing phylogenetic methods to quantify the likelihood of any pathogen evolutionary history at both the intra-host and inter-host levels. Using iSNV data in JUNIPER resulted in an increase in the average posterior probabilities assigned to true transmission links in synthetic outbreaks, as well as an improvement in accuracy over BadTrIP (which uses iSNV data) [10], outbreaker2 [41], and TransPhylo [9]. This improvement was also seen in validation on *in vivo* outbreaks with known or epidemiologically-predicted transmission links, as seen in **Figure 4**.

While JUNIPER incorporates user-provided epidemiological information about the pathogen and outbreak being studied, the resulting reconstructions were robust to variation in these inputs. For each synthetic outbreak used in model validation, we ran JUNIPER once with correctly-specified user inputs and once with default user inputs for the generation interval distribution, sojourn interval distribution, offspring distribution, and other parameters. Across all outbreaks, we observed almost no decrease in transmission inference accuracy (see **Figure 3D–E** and **Supplementary Figure S1K–L**), showing JUNIPER to be accurate even when the user misspecifies various epidemiological quantities. The only scenario in which user input significantly affected reconstruction accuracy was in the South Africa SARS-CoV-2 outbreak, though we emphasize that the lack of a known ground truth in this study makes these accuracies subject to human error. Overall, this finding demonstrates in particular that JUNIPER is effective for analyzing less-studied pathogens, such as novel pathogens, whose epidemiological characteristics have wide uncertainty and hence would not be known to a user.

Beyond its methodological improvements, JUNIPER offers a number of computational benefits compared to existing methods, especially for large datasets. By modeling transmission as a stochastic branching process, JUNIPER avoids computing the coalescent prior, allowing for seamless parallelization over subtrees. Moreover, as discussed in **Methods**, JUNIPER explicitly represents the time of each mutation event in an outbreak. As a result, the cost of computing the evolutionary probability for any phylogenetic tree produced by JUNIPER is far cheaper than for existing methods, such as BEAST/BEAST2––resulting in a decreased runtime despite an increased parameter space dimension. Finally, JUNIPER incorporates numerous novel MCMC moves that accelerate convergence by explicitly proposing configurations with fewer mutations and hence lower evolutionary cost. Together, these features make population-scale transmission inference possible.

We envision several areas of future development for JUNIPER. First, the assumption that bottlenecks are complete limits JUNIPER to pathogens known to exhibit narrow bottlenecks. While JUNIPER does offer a setting that allows incomplete bottlenecks, it requires additional assumptions and approximations, which we hope to refine in subsequent work. JUNIPER further makes the approximation that due to the rapid exponential growth of the within-host population size post-inoculation, all coalescences occur at the time of infection. This assumption prohibits multiple subsequent coalescences occurring within a host, replacing them with a single multifurcation event, and may hence introduce mild biases to the inferred phylogenetic tree and evolutionary parameters. This issue could be resolved by adopting a multispecies coalescent model, though the implementation would likely be far less efficient than JUNIPER in its current form. JUNIPER also assumes the relatively simplistic Jukes-Cantor model of evolution and a stochastic-exponential effective population size. We make these assumptions for a proof-of-concept, but they could easily be relaxed using standard methods from evolutionary theory, e.g. a HKY substitution model [45] and/or a Skygrid model [46] for the effective population size. We plan to implement these additional models as JUNIPER modules in the near future. Lastly, JUNIPER is currently implemented in R; using a compiled language such as C++ would offer a considerable speedup.

Beyond methodological improvements to JUNIPER itself, the analyses produced by JUNIPER in combination with epidemiological metadata could be made more robust with the availability of information on the data collection process. For instance, in the H5N1 case study, our results on transmission rates by state (**Figure 5B**) may differ if certain states investigated, sampled, and sequenced animals colocated with known positive cases, while other states did not. Geographic data at the county or farm level, rather than only state, could further elucidate possible discrepancies in sampling. Similar reasoning applies to our results on transmission rates by vaccination status in the Massachusetts SARS-CoV-2 case study, which may differ in the presence of a correlation between vaccination status and probability of sampling (**Figure 5E–F**). Nonetheless, in the absence of a known sampling scheme, the assumption that samples are collected independently with equal, unknown sampling probability offers a reasonable baseline.

Given JUNIPER’s versatility in handling genomic-epidemiological datasets, and with increased availability of data and continued methodological development, we hope JUNIPER can help make accurate real-time transmission reconstruction a mainstay of infectious disease analysis and response.

## METHODS

Here, we describe our joint likelihood function for transmission networks, phylogenetic trees, and intrahost variants, which together make up the Bayesian statistical model behind JUNIPER. Specifically, “transmission network” refers to the tree of all host-to-host pairs, the times at which each host was infected, and the times at which each host was sampled (if they were sampled); and “phylogenetic tree” refers not only to the sequence of coalescences among lineages observed in the outbreak, but also the times at which mutation events occurred. Explicitly modeling mutation times is somewhat uncommon in the Bayesian phylogenetics literature, but it has been shown to afford major computational advantages [36], and in particular it makes our within-host evolutionary model tractable.

### Transmission Model

We model transmission networks as random branching processes with randomly-drawn generation intervals. Specifically, we model the number of offspring (number of cases infected by a given case) as Negative Binomial, which is an overdispersed probability distribution and hence can capture superspreading. Generation intervals are taken to be Gamma-distributed, a distribution that allows enough flexibility in terms of its mean and variance to capture a wide range of pathogen dynamics. Given a transmission network, we model each host being sampled (i.e., tested and sequenced, and hence part of our dataset) with some probability π, independently across hosts. Finally, we model the time period between a sampled host being infected and tested as again being Gamma-distributed, for analogous reasons. This transmission and sampling model is identical to that of TransPhylo [37]––the central difference being that JUNIPER infers both phylogenies and transmission networks, whereas TransPhylo treats the phylogeny as fixed.

To conduct Bayesian inference using this model, we must compute the probability that a random epidemic generated according to the above process *contains* a proposed transmission network configuration, as well as possibly other cases that were never observed. Here, a proposed transmission network refers to the transmission network among the sampled cases, as well as any other unobserved cases that are ancestral to the sampled ones. Per TransPhylo’s terminology, we refer to cases in such a proposed transmission network as *included*, and other cases as *excluded* [37]. In JUNIPER, we explicitly model the times of infection of all included cases. Hence, to compute the desired probability, we integrate the probabilities of all transmission networks containing the included cases. This integral may be computed using a combination of numerical and analytic techniques, as explained by Equations 8–11 of TransPhylo [37].

### Global Evolutionary Model

While the timing and connectivity of a transmission network does not determine the shape of its underlying phylogenetic tree in general, we make two assumptions in JUNIPER that put these structures in one-to-one correspondence. The first is our central assumption that transmission bottlenecks consist of one virion, which ensures that exactly one lineage on the underlying phylogeny infects each host. The second is that all coalescence events within a given host occur at the time of infection for said host. This assumption, analogous to one made in the recent work of Stolz et al. [25], follows from the fact that within-host viral replication immediately following infection exhibits rapid exponential growth, making earlier coalescent events exponentially more likely than later ones [47].

Given a phylogenetic tree topology, we model molecular evolution as a continuous-time Markov process, in which we parameterize the probability of a given substitution at a given site per unit time. In the simplest such model, known as the Jukes-Cantor model [35], all possible substitutions have the same probability, and substitutions occur independently across different sites on the genome. Under this model, the likelihood associated with a phylogenetic tree and its explicit mutation times is proportional to

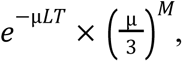

where μ is the mutation rate in substitutions per site per unit time, *L* is the length of the genome, *T* is the total evolutionary time (sum of all branch lengths on the phylogeny), and *M* is the number of mutations on the phylogeny. While the Jukes-Cantor model gives rise to a particularly simple formula for the phylogenetic tree likelihood, other models (e.g. the HKY model [45]) lead to likelihoods that are just as straightforward to compute; we plan to implement these models in future versions of JUNIPER [36].

### Intrahost Evolutionary Model

Our intrahost evolutionary model arises from the key observation that if we assume negligible genetic drift over the course of a single infection rapid exponential growth of the within-host viral population, and no selective advantage, then the only iSNVs observed in deep sequencing data must have occurred very soon after inoculation. Without selective advantage, a variant that first emerges at the time when the viral population consists of *n* particles will make up roughly 1/*n* of the viral population from that time onwards, and for sufficiently large *n*, 1/*n* will almost certainly be below the limit of detection.

To quantify this phenomenon, we model within-host viral replication as a pure-birth process, parameterized as the number of particles *n*(*t*) at time *t* post-inoculation being equal to exp(β*t*) with β a free parameter. Under a Jukes-Cantor model, the number of virions in the population at the time of the first mutation event at a given site on the genome is approximately distributed as Exponential with mean β/μ (see <u>Supplementary Text, Section A.1</u> for derivation). Then, conditional on a viral population of one particle with a given mutation and *n* particles without it, we model the proportion of deep-sequencing reads exhibiting the mutation as following a Beta(1, *n*) distribution. This model was proposed by Leonard et al. [48], and results from modeling the number of descendants of each virion in a population as independent and identically distributed Exponential random variables.

Combining our result on the viral population size when a new lineage emerges with the Leonard et al. model, we derive a probability density function *f* for the fraction *x* of the viral population that exhibits a within-host variant, given the ratio *r* = μ/β:

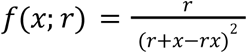

(see <u>Supplementary Text, Section A.2</u> for derivation). This probability density function applies to all iSNVs that are *not* transmitted from one host to another, and hence do not appear as part of the phylogeny linking the cases in our dataset. The explicit-mutation format of this phylogeny tells us the exact time at which each *transmitted* iSNVs first arises, and hence the within-host population size at said time. From there, we may directly apply the Leonard et al. model to obtain the probability distribution of the frequency of the corresponding within-host variant. See <u>Supplementary Text, Section A.5</u> for complete details.

### Implementation

We implement our model using Markov chain Monte Carlo (MCMC) with a custom Metropolis-Hasting sampler. Broadly, our moveset consists of the following operators or combinations thereof:

● Adjust the mutation rate μ.
● Adjust the sampling rate π.
● Adjust the reproductive number *R*.
● Adjust the number and timing of transmissions along a given branch of the phylogenetic tree.
● Adjust the timing of mutation events along a given branch of the phylogenetic tree.
● Adjust the genotype of a given host in the transmission network.
● Adjust the time of infection of a given host.
● Assign a new infector to a given host.
● Add or remove a host from the transmission network.

Moreover, to reduce our state space to a subset of plausible evolutionary histories, we require that the genotype of each unobserved host in the network be *locally parsimonious*, meaning that it induces as few mutations as possible along the branches leading into and out of the host. For instance, if the branch leading to an unobserved host included the C123T substitution, and all branches leading out of the host included the T123C substitution, then the host’s genotype would *not* be locally parsimonious, as the genotype C at site 123 would induce fewer total mutations than T. Configurations that obey are orders of magnitude more likely than those that do not, as the latter incur a penalty on the order of the mutation rate for each additional substitution. For full details of the MCMC implementation, see <u>Supplementary Text, Section B</u>.

### Synthetic Outbreak Generation

To validate JUNIPER, we simulated outbreaks using simulatR, an R package we developed for this purpose. This package first generates transmission networks by starting with a single patient zero and sampling the number of infectees per infectious case as Negative Binomial with mean *R* and overdispersion parameter ψ. We sampled all generation intervals, i.e. the difference in time of infection for a donor-recipient pair, as i.i.d. Gamma random variables with mean and variance μ_g_ and σ_g_^2^, respectively. To select which positive cases appeared in our dataset, we defined two parameters π and ρ_s_, with ρ_s_ optionally being left unspecified (i.e. ρ_s_ = NA). For simulations in which ρ_s_ was left unspecified, we chose which cases are sampled and sequenced as i.i.d. with probability π. If ρ_s_ was specified, we chose whether patient zero is sampled with probability 1/2. The probability that a downstream infectee is sampled and sequenced is ρ_s_ if its infector is sampled and sequenced; otherwise, it is 1 – ρ_s_. Note that in this case, the parameter π is ignored. For sampled and sequenced cases, we sampled sojourn intervals (time from inoculation to sampling) as i.i.d. Gamma random variables with mean and variance μ_s_ and σ ^2^, respectively. We stopped generating each outbreak upon obtaining the first *n*_obs_ = 50 sampled cases, chronologically. We discarded epidemics that ended before generating *n*_obs_ sampled cases.

Having sampled the transmission network, we then generated an underlying phylogenetic tree by assuming all coalescences occur at the time of inoculation, and that each host is inoculated by exactly one lineage. For instance, if host A infects hosts B, C, and D, then the (unique) lineages to infect hosts B, C, and D all coalesce in a single multifurcation event at the time of inoculation of host A. This model is reasonable under the assumption that transmission bottlenecks are complete and that the within-host effective population size grows rapidly immediately after inoculation, which is reasonable for acute viral infections. Given the underlying phylogenetic tree, we sampled mutation events according to the Jukes-Cantor model with a uniform mutation rate across sites on the genome. This means that the number of mutation events at each site along a branch of length *t* follows a Poisson distribution with mean μ*t*; the times of the mutations are uniformly distributed over the duration of the branch; and the new nucleotide at the given site is selected uniformly at random from the three possible choices of different nucleotide.

We generate within-host variants by sampling the earliest time post-inoculation that a mutation occurs at each site on the genome within each host, according to the Jukes-Cantor model. Note that for some sites, the earliest time of mutation may have already been sampled in the previous step (see <u>Supplementary Text, Section A.3</u> for detailed treatment of this case, as the within-host model used by JUNIPER is identical to the one we use to simulate within-host data). We model the within-host effective population size at time *t* post-inoculation as *e*^β*t*^. If the first mutation at a given site occurs at time *t*, then the viral population at that time consists of one virion with the mutation and *e*^β*t*^ without it. Following the approach of Leonard et al. [48], we then sample the fraction of mutated particles at the time of sampling as Beta(1, *e*^β*t*^).

Finally, to generate synthetic NGS data, we report the consensus genome and all intrahost variants with minor allele frequency above 3%. We model ambiguous sites by reporting the correct nucleotide call(s) with probability γ and reporting a missing nucleotide call (N) with probability 1 – γ, i.i.d. across sites. We allow the user to specify samplers for read depth and strand bias; in this paper, we simply set all read depths to 10,000 and all strand biases to 0. In JUNIPER, the interpretation of read depth below a user-specified threshold (default: 100) or strand bias above a user-specified threshold (default: 10 on a phred-scale) is the same as a missing nucleotide call, so we need only vary γ to assess robustness to sequencing issues.

### Analysis of Sequencing Data

SARS-CoV-2 sequencing data for the analysis of the within-host variant frequency distribution was downloaded from the NCBI SRA under BioProject PRJNA715749. A subset of these sequences was used for the transmission reconstruction of 13,570 Massachusetts cases. We used LoFreq version 2.1.5 to call intrahost single nucleotide variants (iSNVs). We then filtered out calls with < 100 read depth, < 10 minor allele read count, and *p*-value of Fisher’s exact test for strand bias < 0.05. In addition, as a means of masking highly-variable sites, we compiled a table listing the number of times across the entire dataset that each possible iSNV appeared with a frequency above 3%. From this dataset, we calculated the probability of a randomly chosen iSNV in a randomly chosen host occurring at each site on the viral genome. Based on these probabilities, we masked sites whose probability of being chosen more than once among 1,000 trials exceeded 5%. This method was designed to model the probability of the same iSNV arising *de novo* more than once in an outbreak of 1,000 people.

H5N1 sequencing data was downloaded from the Nextstrain H5N1 North American cattle outbreak build [49,50], which sources data from the USDA, GenBank, and https://github.com/andersen-lab/avian-influenza. Genomes were filtered to include only the cattle H5N1 virus genomes (i.e., H5N1 genomes from this outbreak but sampled from other animals were excluded) with complete sampling collection dates, resulting in 1,639 genomes. The 8 gene segments for each virus were concatenated and aligned against A/cattle/Texas/24-008749-003/2024 using MAFFT v7.525 [51]. We inferred a maximum likelihood tree of the H5N1 genomes using IQ-TREE 2 [38] under a GTR+G4+F substitution model. We then used TempEst [52] to root the tree using least residual squares and removed any genomes with a residual greater than at least 3 standard deviations (away from 0), leaving 1,628 genomes. This sequence data included genomes isolated from milk, tissue samples, nasal swabs, udder swabs, oropharyngeal swabs, and urine. We filtered these 1,628 samples to those with available intrahost variation data as provided by Andersen Lab GitHub repository, leaving the 1,519 sequences examined in this paper.

All other sequencing data used in this study were taken from previously published research: see Johnson et al. [18] for the BRSV dataset and San et al. [43,53] for the South Africa nosocomial SARS-CoV-2 dataset.

## CODE AND DATA AVAILABILITY

JUNIPER is available as an R package at <u>github.com/broadinstitute/juniper0</u>. JUNIPER requires ape [54], ggplot2 [55], extraDistr [56], and TransPhylo [9] as dependencies. Code used to generate the simulated outbreaks is available as a second R package, simulatR, at <u>github.com/broadinstitute/simulatR</u>. simulatR requires ape [54] and ggplot2 [55] as dependencies. Code to produce all figures and analyses in this paper is available at <u>github.com/broadinstitute/juniper-analyses</u>, and requires the following packages: ape [54], ggplot2 [55], igraph [57], readxl [58] reshape2 [59], lubridate [60], coda [61], and cowplot [62].

## ETHICAL APPROVALS

This study was reviewed and approved by the Massachusetts Department of Public Health Institutional Review Board (IRB) under protocol no. 1603078 and conducted at the Broad Institute via a reliance agreement.

## Data Availability

JUNIPER is available as an R package at github.com/broadinstitute/juniper0. JUNIPER requires ape [54], ggplot2 [55], extraDistr [56], and TransPhylo [9] as dependencies. Code used to generate the simulated outbreaks is available as a second R package, simulatR, at github.com/broadinstitute/simulatR. simulatR requires ape [54] and ggplot2 [55] as dependencies. Code to produce all figures and analyses in this paper is available at github.com/broadinstitute/juniper-analyses, and requires the following packages: ape [54], ggplot2 [55], igraph [57], readxl [58] reshape2 [59], lubridate [60], coda [61], and cowplot [62].

https://github.com/broadinstitute/juniper0

https://github.com/broadinstitute/simulatR

https://github.com/broadinstitute/juniper-analyses

## ACKNOWLEDGEMENTS

We graciously acknowledge A.E. Lin for his feedback on the within-host variation model developed in this paper; M. Khan for his work on improving the JUNIPER codebase; F.Y.C. Liu for his feedback on the manuscript; the team at Fathom Information Design for their exploration of transmission network visualizations; and D. Grant, J.D. Sandi, M. Momoh, and the lab at Kenema Government Hospital in Sierra Leone for being early adopters of reconstructR, the predecessor to JUNIPER.

## AUTHOR CONTRIBUTIONS

Conceptualization: I.S., P.V., M.D.M., P.C.S.; Methodology: I.S., P.V., M.D.M.; Software: I.S.; Validation: I.S.; Formal analysis: I.S.; Investigation: I.S., G.K.M., J.E.P.; Resources: I.S., G.K.M., J.E.P., C.M.B., L.C.M., M.B.; Data Curation: C.M.B., L.C.M., M.B.; Writing – Original Draft: I.S.; Writing - Review & Editing: I.S., G.K.M., T.B., L.A.K., B.A.P., S.F.S., P.V., M.D.M., P.C.S.; Visualization: I.S.; Supervision: S.F.S., D.J.P., B.L.M., A.O., P.V., M.D.M., P.C.S.; Project administration: P.V., M.D.M., P.C.S.; Funding acquisition: D.J.P., B.L.M., A.O., P.C.S.

## SUPPLEMENTARY FIGURES

**Supplementary Figure S1:**
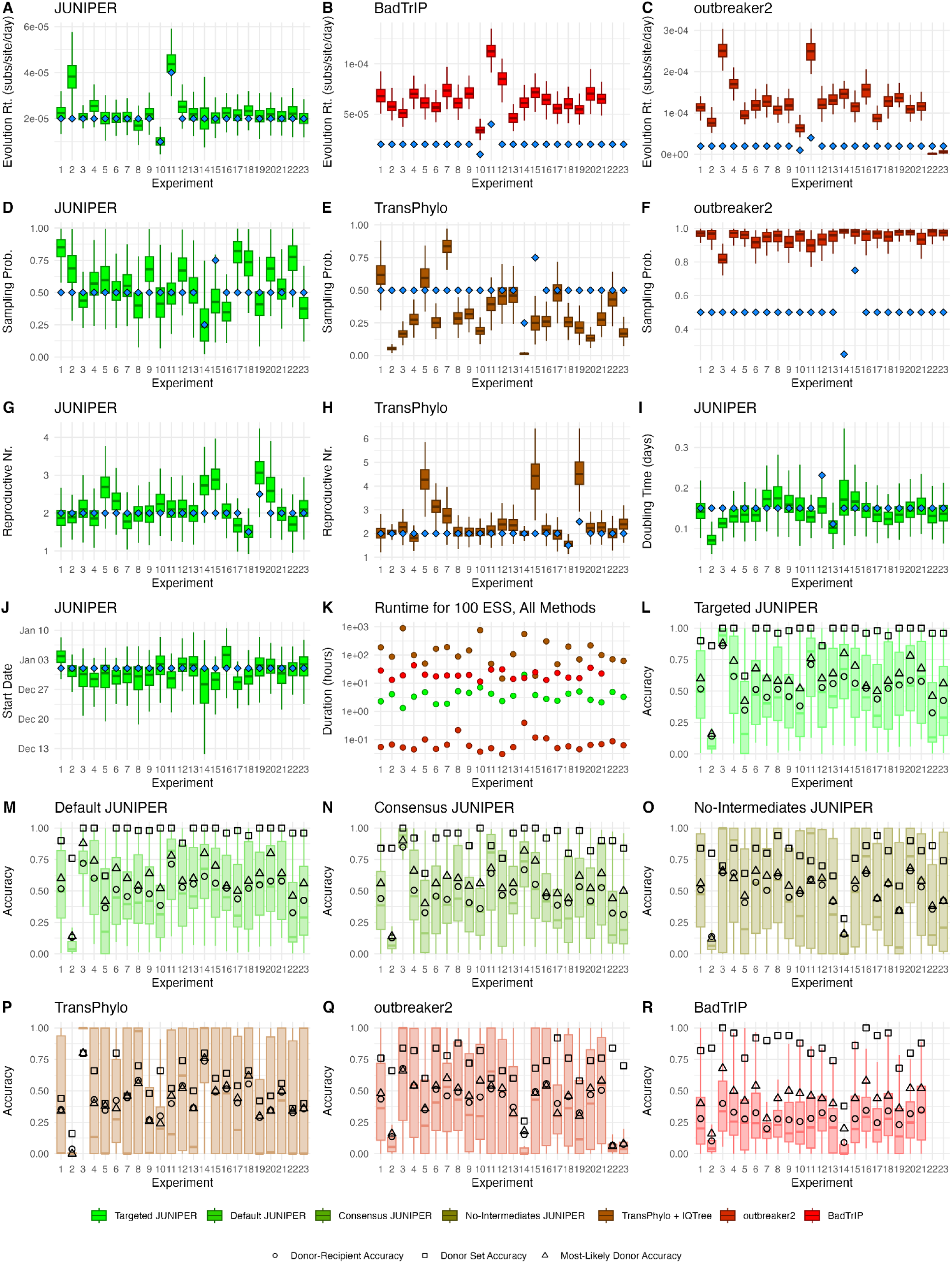
Assessing JUNIPER’s performance on 23 simulated outbreaks. **(A–H)** Posterior density of the **(A–C)** evolution rate, **(D–F)** sampling rate, **(G–H)** reproductive number, **(I)** within-host effective population size doubling time, and **(J)**, inferred by **(A, D, G, I, J)** JUNIPER, **(B)** BadTrIP, **(C, F)** outbreaker2, and **(E, H)** TransPhylo. The true parameter value is shown in blue. **(K)** Runtime to reach an effective sample size of 100 for JUNIPER, TransPhylo, outbreaker2, and BadTrIP. **(L–R)** Distribution of posterior probabilities assigned to ground-truth direct or indirect transmission links (boxplots), donor-recipient accuracy (circles), donor set accuracy (squares), and most likely donor accuracy (triangles) for **(L)** JUNIPER with correctly-specified input parameters, **(M)** JUNIPER with default input parameters, **(N)** JUNIPER without inferring unsampled intermediates, **(O)** JUNIPER without access to within-host variation data, **(P)** IQ-TREE and TransPhylo, **(Q)** outbreaker2, and **(R)** BadTrIP.

**Supplementary Figure S2:**
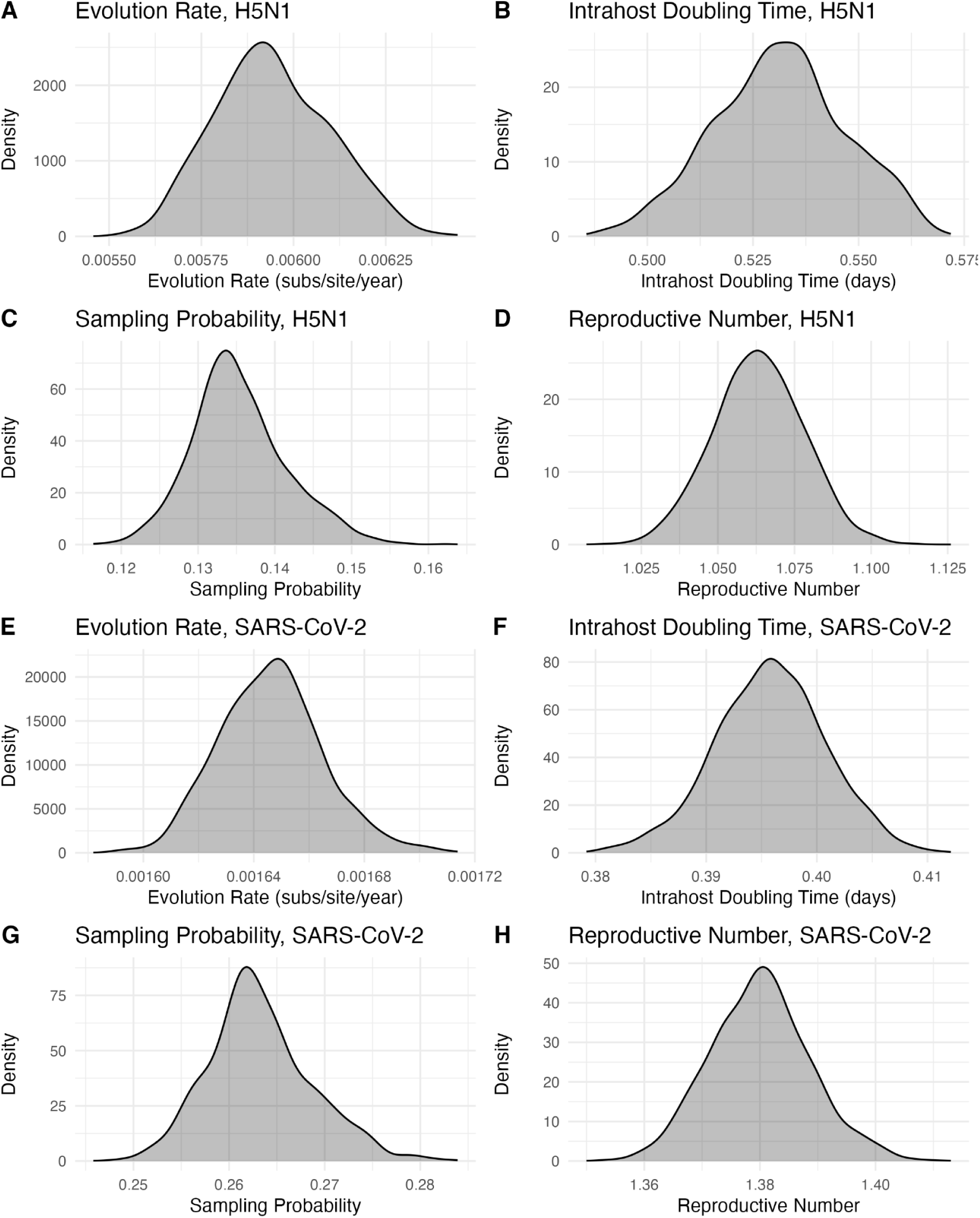
Posterior parameter densities for the bovine H5N1 and Massachusetts SARS-CoV-2 datasets. Posterior densities of the **(A, E)** evolution rate, **(B, F)** doubling time for the within-host effective population size, **(C, G)** sampling probability, and **(D, H)** reproductive number for **(A–D)** H5N1 and **(E–H)** SARS-CoV-2.

**Supplementary Figure S3:**
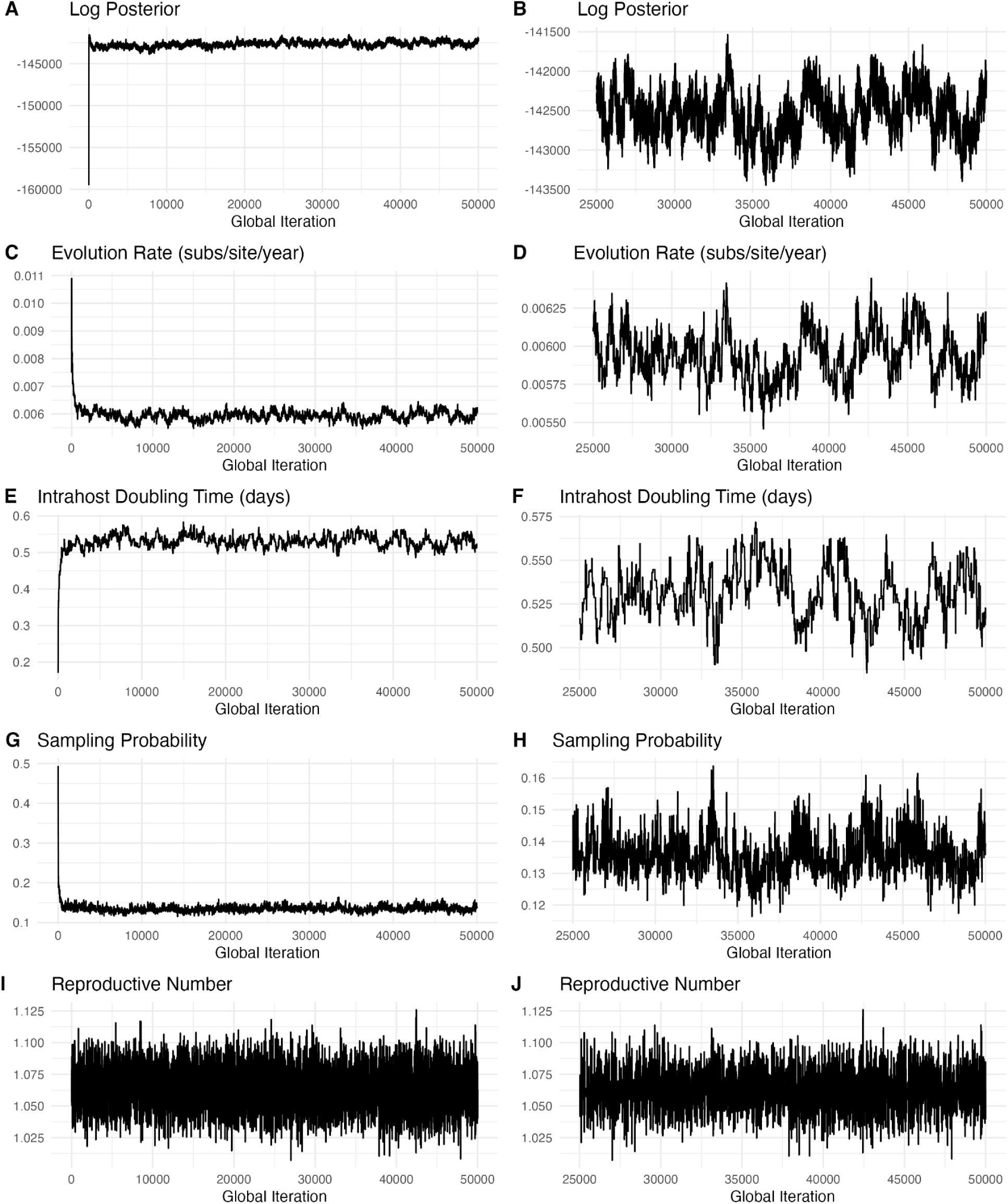
MCMC diagnostics for 1,519 H5N1 cases in cattle. Values of the **(A–B)** log posterior, **(C–D)** evolution rate, **(E–F)** within-host effective population size doubling time, **(G–H)** sampling rate, and **(I–J)** reproductive number for each MCMC iteration. Panels on the right-hand side of the figure show iterations after a 50% burnin period; panels on the left-hand side show all iterations. The effective sample sizes based on each trace post-burnin were 68, 53, 60, 289, and 2,480, respectively.

**Supplementary Figure S4:**
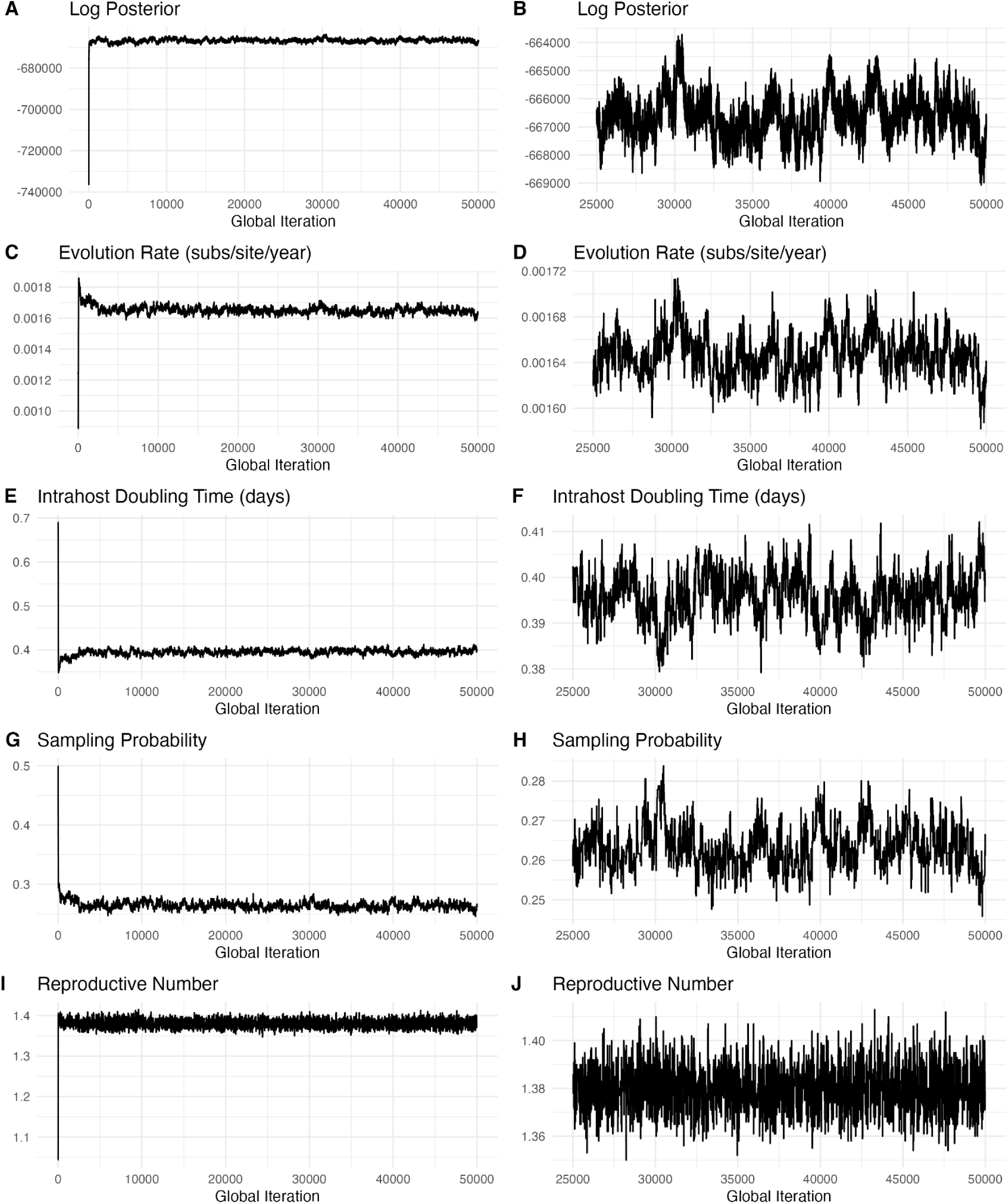
MCMC diagnostics for 13,570 Massachusetts SARS-CoV-2 genomes. Values of the **(A–B)** log posterior, **(C–D)** evolution rate, **(E–F)** within-host effective population size doubling time, **(G–H)** sampling rate, and **(I–J)** reproductive number for each MCMC iteration. Panels on the right-hand side of the figure show iterations after a 50% burnin period; panels on the left-hand side show all iterations. The effective sample sizes based on each trace post-burnin were 82, 117, 139, 173, and 1,534, respectively.

## SUPPLEMENTARY TABLES

**Supplementary Table S1:** Parameters of each experiment. , where μ_g_ and σ_g_^2^ are the mean and variance of the Gamma-distributed generation interval, respectively; μ_s_ and σ_s_^2^ are the mean and variance of the Gamma-distributed sojourn interval, respectively; μ is the evolution rate in substitutions per site per day; β is the growth rate per day of the within-host viral population (i.e. the within-host effective population size *t* days after inoculation is exp(β*t*)); π is the probability that a case is sampled and sequenced; ρ_s_ (if specified) is the probability that a case is sampled and sequenced given that its its infector is sampled and sequenced; the offspring distribution is Negative Binomial with mean *R* and variance *R*/ψ; and γ is the fraction of non-ambiguous sites in sequencing data. Values differing from those used in experiment 1 (i.e. the default) are highlighted in orange. All genomes had a length of 10,000 nucleotides.

| Experiment # | $\mu_g$ | $\sigma_g^2$ | $\mu_s$ | $\sigma_s^2$ | $\mu$ | $\beta$ | $\pi$ | $\rho_s$ | $R$ | $\psi$ | $\gamma$ |
| --- | --- | --- | --- | --- | --- | --- | --- | --- | --- | --- | --- |
| 1 | 5 | 5 | 5 | 5 | 2.00E-05 | ln(100) | 0.5 |  | 2 | 0.5 | 1 |
| 2 | 2.5 | 5 | 5 | 5 | 2.00E-05 | ln(100) | 0.5 |  | 2 | 0.5 | 1 |
| 3 | 10 | 5 | 5 | 5 | 2.00E-05 | ln(100) | 0.5 |  | 2 | 0.5 | 1 |
| 4 | 5 | 2.5 | 5 | 5 | 2.00E-05 | ln(100) | 0.5 |  | 2 | 0.5 | 1 |
| 5 | 5 | 10 | 5 | 5 | 2.00E-05 | ln(100) | 0.5 |  | 2 | 0.5 | 1 |
| 6 | 5 | 5 | 2.5 | 5 | 2.00E-05 | ln(100) | 0.5 |  | 2 | 0.5 | 1 |
| 7 | 5 | 5 | 10 | 5 | 2.00E-05 | ln(100) | 0.5 |  | 2 | 0.5 | 1 |
| 8 | 5 | 5 | 5 | 2.5 | 2.00E-05 | ln(100) | 0.5 |  | 2 | 0.5 | 1 |
| 9 | 5 | 5 | 5 | 10 | 2.00E-05 | ln(100) | 0.5 |  | 2 | 0.5 | 1 |
| 10 | 5 | 5 | 5 | 5 | 1.00E-05 | ln(100) | 0.5 |  | 2 | 0.5 | 1 |
| 11 | 5 | 5 | 5 | 5 | 4.00E-05 | ln(100) | 0.5 |  | 2 | 0.5 | 1 |
| 12 | 5 | 5 | 5 | 5 | 2.00E-05 | ln(20) | 0.5 |  | 2 | 0.5 | 1 |
| 13 | 5 | 5 | 5 | 5 | 2.00E-05 | ln(500) | 0.5 |  | 2 | 0.5 | 1 |
| 14 | 5 | 5 | 5 | 5 | 2.00E-05 | ln(100) | 0.25 |  | 2 | 0.5 | 1 |
| 15 | 5 | 5 | 5 | 5 | 2.00E-05 | ln(100) | 0.75 |  | 2 | 0.5 | 1 |
| 16 | 5 | 5 | 5 | 5 | 2.00E-05 | ln(100) | 0.5 | 0.25 | 2 | 0.5 | 1 |
| 17 | 5 | 5 | 5 | 5 | 2.00E-05 | ln(100) | 0.5 | 0.75 | 2 | 0.5 | 1 |
| 18 | 5 | 5 | 5 | 5 | 2.00E-05 | ln(100) | 0.5 |  | 1.5 | 0.5 | 1 |
| 19 | 5 | 5 | 5 | 5 | 2.00E-05 | ln(100) | 0.5 |  | 2.5 | 0.5 | 1 |
| 20 | 5 | 5 | 5 | 5 | 2.00E-05 | ln(100) | 0.5 |  | 2 | 0.25 | 1 |
| 21 | 5 | 5 | 5 | 5 | 2.00E-05 | ln(100) | 0.5 |  | 2 | 0.75 | 1 |
| 22 | 5 | 5 | 5 | 5 | 2.00E-05 | ln(100) | 0.5 |  | 2 | 0.5 | 0.8 |
| 23 | 5 | 5 | 5 | 5 | 2.00E-05 | ln(100) | 0.5 |  | 2 | 0.5 | 0.9 |

**Supplementary Table S2:**
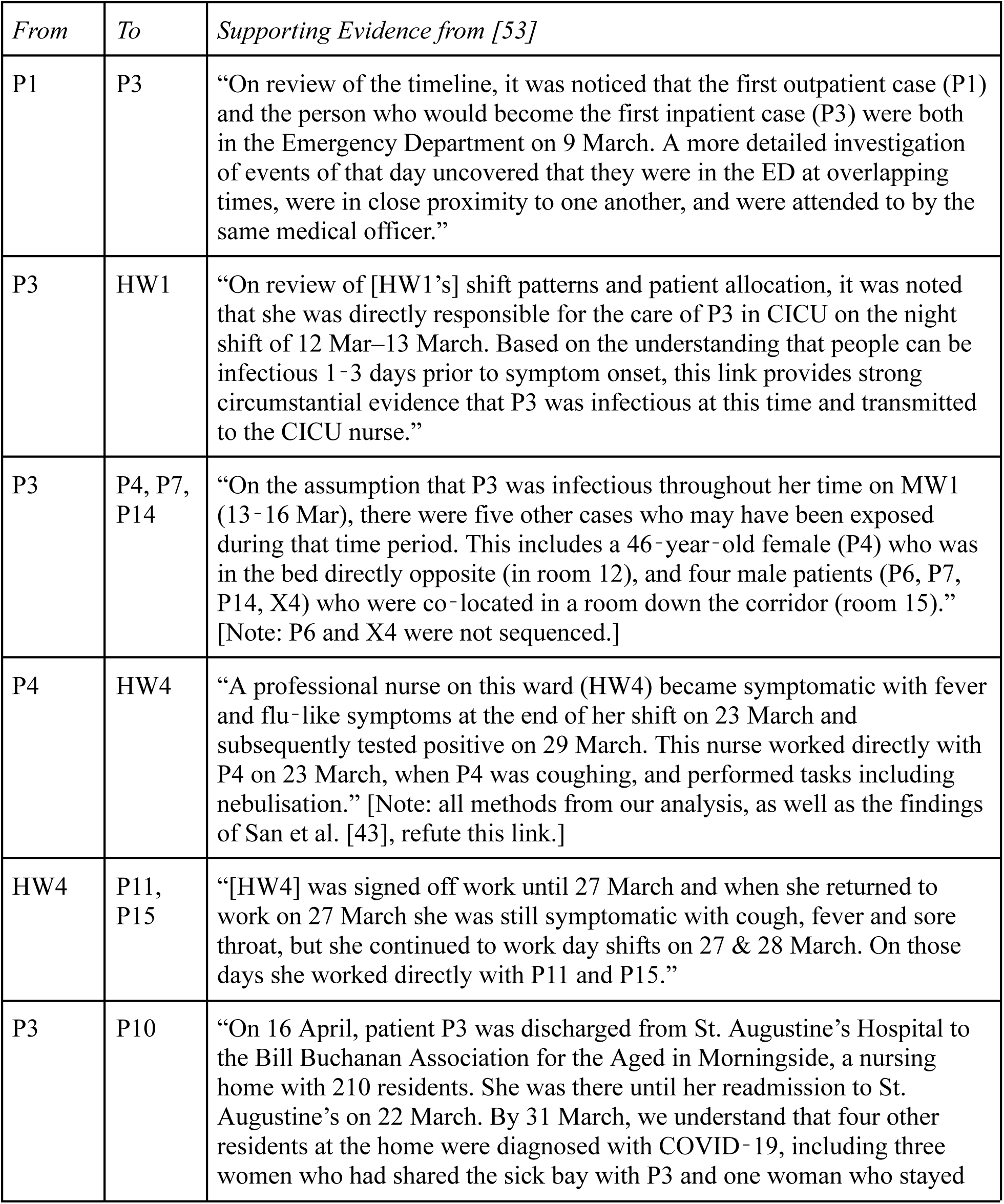
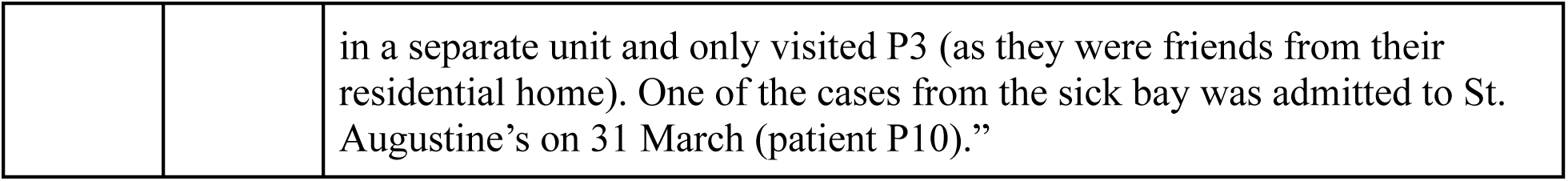
Epidemiological evidence supporting the putative transmission links identified in the South Africa SARS-CoV-2 nosocomial study.

