## Supplementary Text for "JUNIPER: Reconstructing Transmission Events from Next-Generation Sequencing Data at Scale"

#### A Intrahost Evolutionary Model

##### A.1 Viral Population Size at First Mutation Event

To determine the probability density function (PDF) of an iSNV exhibiting a certain frequency, we model within-host viral replication as a pure-birth process in which the the viral population at time $t$ after inoculation is:

$$n(t) := e^{\beta t}$$

for some parameter $\beta$. Our first goal is to compute the total evolutionary time elapsed within the host between the time of inoculation and time $t$ past inoculation. For any $s > 0$ and small $\Delta s > 0$, the approximately $n(s)$ virions to exist in the time interval $(s, s + \Delta s)$ contribute a factor of $n(s)\Delta s$ to the total evolutionary time. Taking the limit as $\Delta s \to 0$, the total evolutionary time at $t$—that is, the total amount of time that all lineages in the population have been extant—is given by

$$N(t) := \int_0^t n(s)ds = \frac{n(t) - 1}{\beta}.$$

Here the $-1$ in the numerator is the constant of integration, chosen such that $N(0) = 0$. Now, under a Jukes Cantor model, the amount of evolutionary time $T$ until the first mutation event at a given site on the genome is distributed as Expo($\mu$), i.e. the Exponential distribution with a mean of $1/\mu$. Setting

$$T = \frac{n(t) - 1}{\beta}$$

and rearranging, we obtain

$$n(t) = 1 + \beta T.$$

Treating $T$ as random, we have that

$$\beta T \sim \text{Expo}\left(\frac{\mu}{\beta}\right)$$

and hence the population size at the time $t$ of the first mutation also approximately follows this distribution, assuming $\mu/\beta$ is small. To simplify notation moving forward, let $r = \mu/\beta$.

#### A.2 Proportion of Mutated Particles

Given $n(t)$, we model the proportion $x$ of viral particles in sequencing data to exhibit the new mutation as $\text{Beta}(1, n(t))$. Hence, the marginal PDF $f$ of this proportion is obtained by the integral

$$f(x;r) = \int_1^\infty f_{\text{Beta}}(x;1,u) f_{\text{Expo}}(u-1;r)\,du$$

where $f_{\text{Beta}}(x;a,b)$ denotes the $\text{Beta}(a,b)$ PDF evaluated at $x$ and $f_{\text{Expo}}(x;\alpha)$ denotes the $\text{Expo}(\alpha)$ PDF evaluated at $x$. While the resulting PDF admits an analytic form, its corresponding CDF (which we will need later) does not. For this reason, we approximate $n(t)$ as following a discrete $\text{Geom}(r)$ distribution, leveraging the fact that $r$ is very close to 0. The above equation then becomes

$$f(x;r) \approx \sum_{k=1}^{\infty} f_{\text{Beta}}(x;1,k) f_{\text{Geom}}(k-1;r) = \frac{r}{(r+x-rx)^2},$$

a highly tractable PDF whose CDF and moments admit analytic forms. In particular, the CDF $F$ is given by

$$F(x;r) := \int_0^x f(t;r)\,dt = \frac{x}{r+x-rx},$$

a function which often arises in computing the likelihood of within-host variation data. Finally, we note that the probability density function of the frequency of a *specific* within-host variant, i.e. a within host variant arising from an `A` to `C` substitution as opposed to a substitution from `A` to any other nucleotide, is given by

$$\frac{r}{3(r+x-rx)^2},$$

assuming the Jukes-Cantor model.

#### A.3 Likelihood of Within-Host Variation Data

While the previous section defines a probability density function for *de novo* within-host variant frequencies, two adjustments must be made when using it to compute the likelihood of NGS data observed in an outbreak. First, in practice, NGS can reliably detect within-host variants that comprise a fraction of the viral population exceeding some threshhold, usually 3% in practice. If variants with frequencies below some threshhold value $B$ are masked, the likelihood $L_{is}(x;r)$ of a specific *de novo* variant within host $i$ at some site $s$ on the viral genome exhibiting frequency $x$ becomes

$$L_{is}(x;r) = \begin{cases} \frac{B}{r+B-rB}, & x < B \\ \frac{r}{3(r+x-rx)^2}, & x \geq B \end{cases}.$$

This function accounts for a lower threshhold on iSNV frequencies by integrating the PDF of $x$ over all undetectable frequencies, resulting in the $x < B$ piece of the piecewise function. It applies to all iSNVs in a given host at positions on the genome that do *not* mutate on the part of the global phylogeny contained within said host, where *global phylogeny* means the phylogenetic tree connecting the bottlenecks of all hosts in our proposed transmission network as described in the *Global Evolutionary Model* subsection of the main text. Now, consider the case that position $s$ exhibits within-host variation in a host $i$, and that the portion of the global phylogeny contained within host $i$ also exhibits a mutation at site $s$. Since the distribution of the frequency of a *de novo* iSNV at site $s$ depends on the population size at the first mutation event at site $s$, we handle this case by conditioning on whether the mutation at site $s$ on the global phylogeny within $i$ is indeed the first to occur at that site within host $i$. Let $t$ be the time at which such a mutation occurs, measured in time units post inoculation of $i$. Take the mutation on the global phylogeny at site $s$ within host $i$ be a substitution from nucleotide $\mathtt{X}$ to nucleotide $\mathtt{Y}$, and let the two nucleotides observed as within-host variants at site $s$ in host $i$ be $\mathtt{X}$ and $\mathtt{Z}$. First consider the case $\mathtt{Y} = \mathtt{Z}$. If the mutation on the globaly phylogeny is the first to occur at site $s$ in host $i$, the likelihood of the frequency $x$ of that mutation follows a $\mathrm{Beta}(1, n(t))$ distribution, as per the previous section. If not, its density function is obtained by restricting population size at the time of first mutation to have support $[0, n(t)]$ and then applying the same Geometric approximation to the Exponential distribution as before:

$$\frac{1}{3}\sum_{k=1}^{n(t)} f_{\mathrm{Beta}}(x;1,k) f_{\mathrm{Geom}}(k-1;r) = \frac{r(n(t)(r-1)x - n(t)r - 1)((r-1)(x-1))^{n(t)} + r}{3(r(-x)+r+x)^2}.$$

While the left-hand side of the above only makes sense when $n(t)$ is an integer, the right-hand side is defined for any real $n(t)$, and hence we allow $n(t)$ to take on continuous values in practice. The case $\mathtt{Y} \neq \mathtt{Z}$ is analogous, except that if the first mutation to occur at site $s$ within host $i$ is the one on the global phylogeny, the probability of observing a within-host mutation from $\mathtt{X}$ to $\mathtt{Z}$ is 0 under our model.

Once again, in the presence of a lower threshhold $B$ for iSNV frequencies, our observed frequencies will either be greater than $B$ or will be reported as being below the limit of detection. In the latter case, as before, the likelihood is computed by integrating the above density function from 0 to $B$, and ignoring the 1/3 term in front because any substitution whose frequency stays below the limit of detection is possible. Again, this integral admits an analytic form:

$$\int_0^B \sum_{k=1}^{n(t)} f_{\mathrm{Beta}}(x;1,k) f_{\mathrm{Geom}}(k-1;r)\,dx = \frac{(1-r)^{n(t)}\left(-r(1-B)^{n(t)+1} - Br + B + r\right) - B}{B(r-1)-r}$$

We are now ready to write down the complete likelihood $L$ of observing a within-host substitution from $\mathtt{X}$ to $\mathtt{Z}$ with frequency $x$ at site $s$ in a host $i$, considering both the limit of detecting for NGS as well as the possibility of a substitution at site $s$ from $\mathtt{X}$ to $\mathtt{Y}$ on the part of the global phylogeny within host $i$. Let $G$ equal 1 if such a substitution occurs on the global phylogeny, and 0, otherwise. Let $S_{\mathrm{Geom}}(k;r)$ denote the $\mathrm{Geom}(r)$ survival function, i.e.

$$S_{\mathrm{Geom}}(k;r) = \sum_{j=k+1}^{\infty} f_{\mathrm{Geom}}(j;r),$$

and let $F_{\mathrm{Beta}}(x;\alpha,\beta)$ denote the $\mathrm{Beta}(\alpha,\beta)$ CDF, i.e.

$$F_{\mathrm{Beta}}(x;\alpha,\beta) = \int_0^x f_{\mathrm{Beta}}(t;\alpha,\beta)\,dt.$$

Then

$$L_{is}(x;r) = \begin{cases} \frac{x}{r+x-rx}, & x < B, G = 0 \\ \frac{r}{3(r+x-rx)^2}, & x \geq B, G = 0 \\ \frac{(1-r)^{n(t)}\left(-r(1-B)^{n(t)+1} - Br + B + r\right) - B}{B(r-1)-r} + S_{\mathrm{Geom}}(n(t);r) F_{\mathrm{Beta}}(x;1,n(t)), & x < B, G = 1 \\ \frac{r(n(t)(r-1)x - n(t)r - 1)((r-1)(x-1))^{n(t)} + r}{3(r(-x)+r+x)^2}, & x \geq B, G = 1, \mathtt{Y} \neq \mathtt{Z} \\ \frac{r(n(t)(r-1)x - n(t)r - 1)((r-1)(x-1))^{n(t)} + r}{3(r(-x)+r+x)^2} + S_{\mathrm{Geom}}(n(t);r) f_{\mathrm{Beta}}(x;1,n(t)), & x \geq B, G = 1, \mathtt{Y} = \mathtt{Z} \end{cases}.$$

Finally, to compute the likelihood $L$ of all within-host variants observed across all hosts, let $x_{is}$ to be the frequency of a *de novo* within-host variant at site $s$ in host $i$. Letting $\mathbf{X}$ denote the collection of all such $x_{is}$ over $i$ and $s$, we define

$$L(\mathbf{X}; r) = \prod_i \prod_s L_{is}(x_{is}; r).$$

Note that if no within-host variation data is available for site $s$ in host $i$, we take $L_{is}(x_{is}; r) = 1$.

#### A.4 Within-Host Likelihood Adjustment for Incomplete Bottlenecks

Recall that our within-host variation model assumes transmission bottlenecks are always complete, i.e. that each host is first infected by a single virion. In this section, we present a modification to the function $L_{is}$ presented above that can incorporate incomplete bottlenecks, though it requires making several additional assumptions and approximations. Modeling incomplete bottlenecks is particularly challenging because it drastically enlarges the space of plausible phylogenetic trees, given a transmission network: even if we maintain the assumption that all coalescences occur at the time of inoculation, we must infer both the *number* of bottleneck particles to inoculate each host as well as *which* is ancestral to each lineage.

Given this challenge, we propose an approximate model under the assumptions that every bottleneck consists of one or two viral particles. Note that even with this assumption, any number of shared polymorphic sites between a donor and recipient are possible. Moreover, we only ever model a host as being inoculated with two virions if (1) the two ends of the branch terminating at said host both represent observed (sampled and sequenced) hosts, (2) those two hosts share at least one polymorphic site with the same two alleles, and (3) adding a second branch of the phylogenetic tree linking the two hosts decreases the total number of mutation events required to realize the genetic diversity observed in both hosts. These conditions are met when, for instance, a donor and recipient have identical consensus genomes, a shared iSNV at one site, and no other polymorphic sites. This scenario may occur if two virions—one with one genotype at the polymorphic site and one with the other—inoculate the recipient. It may also occur if the polymorphism arises *de novo* in both the donor and recipient, but this latter scenario requires two mutation events, as opposed to one in the former.

Consider the case that conditions (1), (2), and (3) are met for a given two hosts $h$ and $i$ with $h$ ancestral to $i$. First, suppose $s$ is a shared polymorphic site for $h$ and $i$. In this case, we model the inoculum of host $i$ as consisting of two virions, one with each of the two shared alleles at site $s$. Hence, we model the fraction of reads in $i$ at site $s$ exhibiting one of the alleles is $\text{Beta}(1, 1)$, which is the same as the uniform distribution on $[0, 1]$. The probability density function of this distribution is 1 on $[0, 1]$ and 0 elsewhere, so its contribution to the within-host evolutionary likelihood is constant. If $s$ is *not* a shared polymorphic site, then we model both particles to infect host $i$ as having the same genotype, and we revert to the previously-defined $L_{is}$ to compute the likelihood for that site.

We also need to account for the fact that there now exists an additional branch on the phylogenetic tree linking $h$ to $i$. This branch needs no additional mutations, since whatever mutation led to the shared polymorphic site has already been accounted for in the within-host evolutionary likelihood function calculated for host $i$. Moreover, under the assumption of rapid exponential growth of the within-host effective population size immediately after inoculation, said mutation must have occurred just after host $h$ was infected (or was already present in the bottleneck of $h$). Since the new branch extends from the time of mutation in $h$ to the time of inoculation of $i$, its length is approximately the difference in times of inoculation of $h$ and $i$—i.e. the same length as the existing branch from $h$ to $i$.

We now have what we need to define our modified within-host likelihood function, which we will call $\tilde{L}(\mathbf{X}; r)$. Let $H_i = 1$ if conditions (1), (2), and (3) hold in $i$, and 0, otherwise. To account for shared polymorphic sites in $h$ and $i$, let

$$\tilde{L}_{is}(x; r) := \begin{cases} 1, & H_i = 1 \text{ and } s \text{ is a shared polymorphic site between } h \text{ and } i \\ L_{is}(x; r), & \text{otherwise} \end{cases}.$$

Then, to account for the additional branch from $h$ to $i$, let $t_h$ and $t_i$ denote the times of inoculation of $h$ and $i$ respectively, and set

$$\tilde{L}(\mathbf{X}; r) = \prod_i [\exp(-\mu(t_i - t_h))]^{H_i} \prod_s \tilde{L}_{is}(x_{is}; r).$$

The term $\exp(-\mu(t_i - t_h))$ is the Jukes-Cantor likelihood of a branch of length $t_i - t_h$ exhibiting zero mutations.

Because of the additional assumptions and approximations required here, we recommend using this optional adjustment to the likelihood function only in scenarios with little to no consensus-level diversity. Note that it only aids in inferring transmission links based on shared polymorphic sites in the absence of consensus-level changes; the instance of a minor allele being transmitted from a donor and reaching fixation in the recipient is already accounted for without any need for this modification.

##### A.5 Complete Posterior Density

Having defined the intrahost evolutionary model, we can now write the complete posterior density from which we sample using MCMC. To do so, we first establish some notation. Let

$$\mathbf{Y} = (\mathbf{G}, \mathbf{Z}, \mathbf{s})$$

denote our data, where:

- $\mathbf{G}$ is an $n_{\text{obs}} \times n_{\text{bases}}$ matrix with entries in {`A, C, G, T`} whose entry $g_{is}$ is the nucleotide at site $s$ on the consensus genome collected from host $i$, where $n_{\text{obs}}$ is the number of observed hosts and $n_{\text{bases}}$ is the length of the viral genome.

- $\mathbf{Z}$ is an $n_{\text{obs}} \times n_{\text{bases}}$ matrix whose entry $z_{is}$ is the proportion of the viral population exhibiting a minor allele at site $s$ in host $i$. Note that in practice, we mask all multiallelic sites, so $z_{is}$ is well-defined. Depending on the contents of the bottleneck infecting host $i$, the *de novo* within-host variant to arise at site $s$ in host $i$ may have frequency $z_{is}$ or $1 - z_{is}$, hence the need to distinguish between $\mathbf{Z}$ defined here and $\mathbf{X}$ as defined in Section A.3.

- $\mathbf{s}$ is a vector of length $n_{\text{obs}}$ whose $i$th entry is the time that a genome was sampled from case $i$. By convention, we set $\max_i s_i = 0$, and all other entries of $\mathbf{s}$ relative to its maximum. As a result, all entries of $\mathbf{s}$ are non-positive.

Next, let

$$\boldsymbol{\theta} = (n, \mathbf{h}, \mathbf{t}, \mathcal{M}, \mathbf{X}, \mu, R, \psi, \beta, \pi, a_g, \lambda_g, a_s, \lambda_s)$$

where:

- $n$ is the total number of hosts in the network.
- $\mathbf{h}$ is the length-$n$ vector of ancestors, whose $i$th entry $h_i$ is equal to the infector of a host $i$. By convention, the index $i = 1$ denotes the root of the cluster, and we set $h_1$ to be undefined (`NA` in computer terms).
- $\mathbf{t}$ is a vector of length $n$ whose $i$th entry $t_i$ is the time at which host $i$ becomes infected.
- $\mathcal{M}$ is a list of all mutations to occur on the global phylogeny. We organize $\mathcal{M}$ as a list of lists, where the $i$th entry enumerates the mutations along the branch of the phylogeny starting at the bottleneck of host $h_i$ and ending at the bottleneck of host $i$. Each mutation consists of the nucleotide being mutated away from, the position on the genome of the mutation, the nucleotide being mutated into, and the time at which the mutation occurs e.g. `C123T` at time $-10$. Let $|\mathcal{M}|$ denote the total number of mutation events in all of $\mathcal{M}$.
- $\mathbf{X}$ is an $n_{\text{obs}} \times n_{\text{bases}}$ matrix whose entry $x_{is}$ is the proportion of the viral population exhibiting a *de novo* within-host variant site $s$ in host $i$. Depending on the contents of the bottleneck infecting $i$ (which may be deduced from $\mathcal{M}$), $x_{is}$ is equal to either $z_{is}$ or $1 - z_{is}$.
- $\mu$ is the mutation rate, in substitutions per site per unit time.
- $R$ is the reproductive number, i.e. the mean of the offspring distribution.
- $\psi$ is the second parameter of the offspring distribution, which is modeled as Negative Binomial with parameters $\frac{R\psi}{1-\psi}$ and $\psi$ (to ensure the mean equals $R$).
- $\beta$ is the within-host effective population size growth rate, as defined in Section A.1.
- $\pi$ is the probability that a host is sampled.
- $a_g$ is the shape parameter of the generation interval, which we assume to follow a Gamma distribution.
- $\lambda_g$ is the rate parameter of the generation interval, which we assume to follow a Gamma distribution.
- $a_s$ is the shape parameter of the sojourn interval (the time between inoculation and sampling), which we assume to follow a Gamma distribution.
- $\lambda_s$ is the rate parameter of the sojourn interval, which we assume to follow a Gamma distribution.

Using the definitions of $\mathbf{Y}$ and $\boldsymbol{\theta}$, the likelihood $\pi(\mathbf{Y}|\boldsymbol{\theta})$ is given by:

$$\pi(\mathbf{Y}|\boldsymbol{\theta}) = \prod_{i=1}^{n} \Bigg[ (1-\pi_{t_i})^{\mathbb{1}(i \notin \text{obs})} (\pi_{t_i} f_{\text{Gamma}}(s_i - t_i; a_s, \lambda_s)^{\mathbb{1}(i \in \text{obs})}) \alpha_{t_i}(d_i) \\ \times \prod_{j: h_j = i} f_{\text{Gamma}}(t_j - t_i; a_g, \lambda_g) \Bigg] \times \\ \exp\left( -\mu n_{\text{bases}} \sum_{i=1}^{n} \sum_{j: h_j = i} (t_j - t_i) \right) \times \left(\frac{\mu}{3}\right)^{|\mathcal{M}|} \times \\ L(\mathbf{X}; \mu/\beta),$$

where:

- $\mathbb{1}(A)$ is the indicator function of an event $A$.
- obs is the set of observed (i.e. sampled and sequenced) hosts.
- $f_{\text{Gamma}}(x; a, \lambda)$ is the probability density function of a Gamma distribution with shape parameter $a$ and rate parameter $\lambda$ evaluated at $x$.
- $\pi_{t_i} = \pi \int_{t_i}^{0} f_{\text{Gamma}}(x; a_s, \lambda_s)\, dx$ is the probability that host $i$ is sampled by the time 0, i.e. the time of the last sample collection and hence the time at which data collection ends.
- $d_i$ is the number of people infected by host $i$, i.e. the cardinality $|\{j : h_j = i\}|$.
- $\alpha_{t_i}(d_i) = \sum_{k=d_i}^{\infty} \binom{k}{d_i} f_{\text{NBin}}(k; \frac{R\psi}{1-\psi}; \psi)\bar{\omega}_{t_i}^{k-d_i}$, where
  - $f_{\text{NBin}}(k; r, p)$ is the probability mass function of the Negative Binomial distribution with parameters $r$ and $p$ evaluated at $k$, i.e.

$$f_{\text{NBin}}(k; r, p) = \frac{\Gamma(k + r)}{\Gamma(r)k!} p^r (1 - p)^k.$$

  - $\bar{\omega}_{t_i}^{k-d_i}$ is the probability that $k - d_i$ of the offspring of $i$ are unsampled and have no sampled descendants by time 0. This quantity may be calculated numerically using equation 9 of `TransPhylo` (Didelot et al. 2017).
- $L(\mathbf{X}; \mu/\beta)$ is defined in Section A.3. It may optionally be replaced by $\tilde{L}(\mathbf{X}; \mu/\beta)$ as defined in Section A.4.

The first two lines of the likelihood $\pi(\mathbf{Y}|\boldsymbol{\theta})$ and all associated definitions, which account for the probability associated with the transmission network, are adapted from `TransPhylo` (Didelot et al. 2017), equations 8–11. The third line is the Jukes-Cantor model for an explicit mutation representation of a phylogeny (Jukes & Cantor 1969). The final line is the within-host variant frequency model developed in this paper.

Additionally, there are several conditions $\boldsymbol{\theta}$ must satisfy, else we set the value of $\pi(\mathbf{Y}|\boldsymbol{\theta})$ to 0. These conditions are:

- If a position $s$ on the genome in host $i$ is observed, meaning that $g_{is} \in$ {`A, C, G, T`} (as opposed to `N` or `-` or another designator of missing data), then we require that either (a) if no iSNV is observed at site $s$ in host $i$, the bottleneck infecting $i$ must have allele $g_{is}$ at site $s$, or (b) if an iSNV is observed at site $s$ in host $i$, the bottleneck infecting $i$ must have one of the two alleles observed at site $s$.
- The global phylogeny must obey *local parsimony*. This means that for each host $i$, the genotype of the bottleneck infecting host $i$ must be selected to minimize the number of mutations among the portion of the global phylogeny connecting host $i$ to host $h_i$ and hosts $j$ such that $h_j = i$, subject to all bottlenecks in the global phylogeny satisfying the previous condition.

Finally, we must define our prior on $\boldsymbol{\theta}$. In accordance with `TransPhylo` (Didelot et al. 2017), the only non-uniform prior we assign is $R \sim \text{Expo}(1)$, i.e.

$$\pi(\boldsymbol{\theta}) = \exp(-R).$$

Having defined our prior and likelihood, we may now compute the posterior $\pi(\boldsymbol{\theta}|\mathbf{Y})$ up to a constant of proportionality via Bayes' Theorem:

$$\pi(\boldsymbol{\theta}|\mathbf{Y}) \propto \pi(\mathbf{Y}|\boldsymbol{\theta})\pi(\boldsymbol{\theta}).$$

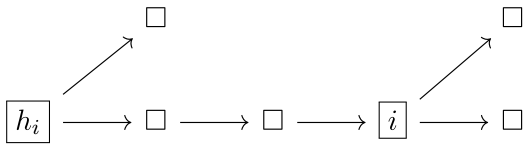


Figure 1: Let boxes represent hosts and arrows represent transmissions. Here, $h_i$ and $i$ are explicit hosts, and the two unlabeled hosts along the transmission chain from $h_i$ to $i$ are referred as the implicit hosts leading to $i$.

### B MCMC Implementation

#### B.1 Moves

We draw samples from the joint posterior distribution of transmission networks, phylogenies, and their underlying parameters using a Metropolis-Hastings sampler with a custom moveset. We separate our MCMC moves into two categories: *global moves*, which affect every node's contribution to the likelihood function, and *local moves*, which only affect the contributions of a few nodes. The global moves are:

1. Adjust the value of the mutation rate $\mu$ by adding a Normal random variable with mean 0 and standard deviation $\mu_0/10$, where $\mu_0$ is a user-specified initial guess of the mutation rate (defaults to $\mu_0 = 2 \times 10^{-5}$).

2. Adjust the value of the sampling rate $\pi$ by adding a Normal random variable with mean 0 and standard deviation 0.05.

3. Adjust the value of the reproductive number $R$ by adding a Normal random variable with mean 0 and standard deviation 0.1.

All of these moves require updating the likelihood function associated with the global phylogeny, the likelihood of the transmission network, and the likelihood function of the within-host evolution at every host. As such, they are relatively expensive, in contrast to the local moves to follow. Before enumerating them, we establish some notation: let an *observed* host refer to a host that is sampled and sequenced, and let an *unobserved* host refer any other host. Let an *explicit* host refer to any host that is either (a) observed, (b) has at least two offspring, or (c) is the root, i.e. the first case to be infected. For an explicit host $i$, we use the notation $h_i$ to mean the most recent explicit host that is ancestral to $i$. Let the *implicit hosts leading to $i$* refer to the non-explicit hosts along the transmission chain from $h_i$ to $i$ (noting that the set of implicit hosts leading to $i$ may be empty). See Figure 1.

For an explicit host $i$, let $\mathbf{t}_i$ denote the times of inoculation for $i$ and the implicit hosts leading to $i$, in decreasing order. For an observed host $i$, let $s_i$ denote the time of sampling. Throughout, we use the notation $\mathbf{x}[k]$ to denote the $k$th element of the vector $\mathbf{x}$, with 1-based indexing. Let $a_g$ and $\lambda_g$ be the shape and rate parameters of the Gamma-distributed generation interval; let $a_s$ and $\lambda_s$ be the shape and rate parameters of the Gamma-distributed sojourn interval. For ease of notation, let $\mu_g$ and $\mu_s$ denote the means of the generation and sojourn intervals, respectively.

Using this notation, we may define our local moves. As some of them are quite complex and notation-heavy, we provide a simple descriptor of each move in the list below.

4. Resample the times of infection along a branch.

5. Resample the times of infection and mutation along a branch.

6. Resample the times of infection for a host $i$ and all hosts along the branches connected to $i$.

7. Resample the times of infection for a host $i$, its ancestors, and all hosts along the branches connected to $i$ and its ancestors.

8. Update the genotype of a host $i$.

9. Resample the ancestor $h_i$ of $i$ to be either the ancestor or an offspring of $h_i$.

10. Resample the ancestor $h_i$ of $i$ to be anyone infected before $i$.

11. Resample the ancestor $h_i$ of $i$ to be anyone infected before $i$, but bias the choice such that hosts with genotypic similarities to $i$ are proposed more often.

12. Pick a pair of hosts $i, j$ where $i$ infects $j$, and rearrange the transmission network such that $j$ infects $i$.

13. Do the same as the previous move, except also set the offspring of $i$ to be infected by $j$, and set the offspring of $j$ to be infected by $i$.

For the remaining three moves, upon choosing to perform the move, we will randomly select one of two possible sub-moves, to ensure reversibility:

15. With probability 1/2, pick a host $h$ and create a new unobserved host $i$ that is infected by $h$. Then, take some of the offspring of $h$ and make them into offspring of $i$. With probability 1/2, delete an unobserved host $i$, and update the ancestor of $i$'s offspring to be $h_i$.

16. With probability 1/2, pick a host $j$ and create a new unobserved host $i$ that infects $j$. Then, take some of the offspring of $j$ and make them into offspring of $i$. With probability 1/2, delete an unobserved host $i$, and update the ancestor of $i$'s offspring to be $j$, one of the former offspring of $i$.

17. With probability 1/2, pick a mutation that occurs twice on the global phylogeny, create a new unobserved host $i$ with this mutation, and rearrange the phylogeny such that this mutation occurs only once. With probability 1/2, delete an unobserved host $i$ and resample the ancestors of the former children of $i$ from the descendants of $h_i$.

The overviews of Moves 4–16 provided above may be implemented as follows, respectively:

4. Select an explicit host $i$ uniformly at random. Resample the number of implicit hosts leading to $i$ as being equal to the rounded value of $(\mathbf{t}_i[1] - \mathbf{t}_{h_i}[1])\mu_g$ with probability 0.95, or drawn from the $\text{Pois}((\mathbf{t}_i[1] - \mathbf{t}_{h_i}[1])\mu_g)$ distribution with probability 0.05. Then, resample $\mathbf{t}_i$ as the cumulative sum of a $\text{Dirichlet}(\mu_g, \mu_g, \ldots, \mu_g)$ draw, rescaled to the interval $(\mathbf{t}_{h_i}[1], \mathbf{t}_i[1])$, and in reverse order.

5. Perform Move 4. Then, resample the times of mutation events along the branch from $\mathbf{t}_{h_i}[1]$ to $\mathbf{t}_i[1]$ as i.i.d. $\text{Unif}(\mathbf{t}_{h_i}[1], \mathbf{t}_i[1])$ draws.

6. Select an explicit host $i$ uniformly at random. Let $J = \{j : h_j = i\}$. Let $t_{\max} = \min\{\{\mathbf{t}_j[1] : j \in J\} \cup s_i\}$. If $i$ is not the root, resample $\mathbf{t}_i[1] \sim \text{Unif}(\mathbf{t}_{h_i}[1], t_{\max})$, and then apply Move 5 to $i$ and to each $j$. If $i$ is the root, let $T$ denote the total evolutionary time, i.e. the difference in time between $\max_j s_j$ and $\mathbf{t}_1[1]$. Let $\mu_\Delta = T/10$ and let
$$p = \frac{\mu_\Delta}{\mu_\Delta + t_{\max} - \mathbf{t}_i[1]}.$$
Then sample
$$\Delta \sim \begin{cases} -\text{Expo}(\mu_\Delta) & \text{with probability } p \\ \text{Unif}(0, t_{\max} - \mathbf{t}_i[1]) & \text{with probability } 1-p \end{cases}.$$
Finally, apply Move 5 to each $j \in J$.

7. Select an explicit host $i$ uniformly at random. Let $J = \{j : h_j = i\}$. Let $I$ denote the set of all explicit hosts ancestral to $i$, including $i$—that is, if we apply the function $i \mapsto h_i$ recursively, we enumerate the elements of $I$. Let $J = \{j : h_j \in I\} \setminus I$. Let
$$\Delta_{\max} = \min\{\{\mathbf{t}_j[1] - \mathbf{t}_{h_j}[1] : j \in J\} \cup \{s_{i'} - \mathbf{t}_{i'}[1] : i' \in I\}\}.$$
As in Move 6, let $T$ denote the total evolutionary time, let $\mu_\Delta = T/10$, and let
$$p = \frac{\mu_\Delta}{\mu_\Delta + \Delta_{\max}}.$$
Then sample
$$\Delta \sim \begin{cases} -\text{Expo}(\mu_\Delta) & \text{with probability } p \\ \text{Unif}(0, \Delta_{\max}) & \text{with probability } 1-p \end{cases}.$$
Set $\mathbf{t}_{i'} \leftarrow \mathbf{t}_{i'} + \Delta$ for each $i' \in I$, and finally, apply Move 5 to each $j \in J$.

8. Select an explicit host $i$ uniformly at random. Let $J = \{j : h_j = i\}$. Resample the genotype of $i$ uniformly at random from the set of genotypes that minimize the number of mutations along the portion of the phylogeny connecting $i$ to each $j \in J$ and to $h_i$, noting that if $i$ is observed, than the genotype may only be updated at sites with missing data and sites with iSNVs. Then resample the times of the mutations on the branch of the phylogeny leading into $i$ uniformly at random. Do the same thing for each $j \in J$.

9. Do one of the following, with equal probability:

   (a) Select an explicit host $i$ uniformly at random. Let $J = \{j : h_j = i\}$. Select $j \in J$ uniformly at random, and set $h_i \leftarrow j$. Then apply Move 4 to $i$. Finally, apply Move 8 to $i$.

   (b) Select an explicit host $i$ uniformly at random. Set $h_i \leftarrow h_{h_i}$. Then apply Move 4 to $i$. Finally, apply Move 8 to $i$.

10. Select an explicit host $i$ uniformly at random. Let $A = \{a : \mathbf{t}_a[1] < \mathbf{t}_i[1]\}$, and sample $h_i$ uniformly from $A$. Then apply Move 4 to $i$. Finally, apply Move 8 to $i$.

11. Select an explicit host $i$ uniformly at random, and let $A$ be as above. For each $a \in A$, let $m_a$ be the number of mutations on the branch of the phylogeny leading into $a$ that also appear on the branch of the phylogeny leading into $i$. Sample $a \in A$ with probability
$$\frac{\exp(m_a/\tau)}{\sum_{a' \in A} \exp(m_{a'}/\tau)}$$
with $\tau = 0.2$. Set $h_i \leftarrow a$; apply Move 4 to $i$; and, finally, apply Move 8 to $i$.

12. Select an explicit host $j$ uniformly at random subject to the condition that $h_j$ and $h_{h_j}$ both exist. Let $i = h_j$ and let $h = h_i$. Set $h_j \leftarrow h$, $h_i \leftarrow j$, and swap the vectors $\mathbf{t}_j$ and $\mathbf{t}_i$. Let $K = \{k : h_k \in \{h, i, j\}\}$. For each $k \in K$, resample the times of mutations on the branch of the phylogenetic tree leading into $k$.

13. Perform Move 12, except after swapping the vectors $\mathbf{t}_j$ and $\mathbf{t}_i$, perform the following additional step: let $K_i = \{k : h_k = i\}$; let $K_j = \{k : h_k = j\}$, set $h_k = i$ for each $k \in K_j$; and set $h_k = j$ for each $k \in K_i$.

14. Do one of the following, with equal probability:

    (a) Select an explicit host $h$ with probability proportional to its degree (i.e. the size of the set $J = \{j : h_j = h\}$), subject to the condition that the degree of $h$ is at least 2. Let $J_0 = \{j : h_j = h\}$. Sample $n_j \sim \text{Unif}(\{2, 3, \ldots, |J_0|\})$ if $h$ is observed, or $n_j \sim \text{Unif}(\{2, 3, \ldots, |J_0| - 1\})$ if $h$ is unobserved. Sample a subset $J$ uniformly at random from the set of subsets of $J_0$ that have cardinality $n_j$. Create a new host explicit host $i$, set $h_i = h$, and set $h_j = i$ for each $j \in J$. Draw $\mathbf{t}_i[1] \sim \text{Unif}(\mathbf{t}_h[1], \min_{j \in J} \mathbf{t}_j[1])$, then draw the rest of the vector $\mathbf{t}_i$ by applying Move 4 to $i$. Then apply Move 4 to each $j \in J$. Finally, apply Move 8 to $i$.

    (b) Select an unobserved explicit host $i$ uniformly at random. Let $J = \{j : h_j = i\}$, and let $h = h_i$. Delete host $i$ and set $h_j = h$ for each $j$ in $J$. Finally, apply Move 5 to each $j \in J$.

15. Do one of the following, with equal probability:

    (a) Select an explicit host $j_1$ uniformly at random. Let $K_0 = \{k : h_k = j_1\}$ and let $h = h_{j_1}$. Sample $n_2 \sim \text{Unif}(\{1, 2, \ldots, |K_0|\})$ if $j_1$ is observed, or $n_j \sim \text{Unif}(\{1, 2, \ldots, |K_0| - 2\})$ if $j_1$ is unobserved. Sample a subset $K$ uniformly at random from the set of subsets of $K_0$ that have cardinality $n_j$. For each $k \in K$, set $h_k = h$. Let $J = K \cup \{j_1\}$. Create a new host explicit host $i$, set $h_i = h$, and set $h_j = i$ for each $j \in J$. Draw $\mathbf{t}_i[1] \sim \text{Unif}(\mathbf{t}_h[1], \min_{j \in J} \mathbf{t}_j[1])$, then draw the rest of the vector $\mathbf{t}_i$ by applying Move 4 to $i$. Then apply Move 4 to each $j \in J$. Finally, apply Move 8 to $i$.

    (b) Select an unobserved explicit host $i$ uniformly at random. Let $J = \{j : h_j = i\}$, let $j_1 = \arg\min_{j \in J} \mathbf{t}_j[1]$, let $K = J \setminus \{j_1\}$, and let $h = h_i$. Delete host $i$, set $h_{j_1} = h$, and set $h_k = j_1$ for each $k \in K$. Finally, apply Move 5 to each $j \in J$.

16. Do one of the following, with equal probability:

    (a) Let $P$ denote the set of positions on the genome that mutate at least once on the global phylogeny. For a position $p \in P$, let $n_p$ be one less than the number of times a mutation at position $p$ occurs on the global phylogeny. Let $B \sim \text{Bernoulli}(0.95)$. If $B = 1$, and if the $n_p$'s are not all equal to 0, sample a position $p$ with probability proportional to $n_p$. Let $J_0$ be the set of explicit hosts $j$ such that the branch of the global phylogeny leading into $j$ has a mutation at position $p$, and note that $|J_0| \geq 2$ by construction of $n_p$. Sample a subset $J$ of cardinality 2 from $J_0$ uniformly at random. Let $h$ be the most recent common ancestor of the two elements of $J$, i.e. the latest-to-be-infected explicit host that is ancestral to both elements of $J$. Create a new host explicit host $i$, set $h_i = h$, and set $h_j = i$ for each $j \in J$. Draw $\mathbf{t}_i[1] \sim \text{Unif}(\mathbf{t}_h[1], \min_{j \in J} \mathbf{t}_j[1])$, then draw the rest of the vector $\mathbf{t}_i$ by applying Move 4 to $i$. Then apply Move 4 to each $j \in J$. Finally, apply Move 8 to $i$.

(b) Select an unobserved explicit host $i$ uniformly at random, subject to the condition that the set $J = \{j : h_j = i\}$ has cardinality 2. Let $h = h_i$. Delete host $i$ and set $h_j = h$ for each $j$ in $J$. For each $j$ in $J$, repeatedly perform the following sequence of actions until it terminates: (1) set $K = \{k : h_k = h_j, k \neq j\}$; (2): with probability $\frac{1}{K+1}$, terminate, or with probability $\frac{K}{K+1}$, select a $k$ uniformly at random from $K$, and set $h_j = k$. Note that repeating these actions must eventually terminate because each iteration takes $j$ one step further from the root, and the entire transmission network can only have finitely many hosts. Finally, apply Move 5 to each $j \in J$.

Note also that to ensure we maintain the property that an unobserved explicit host must either have degree 2 or be the root, we automatically reject moves that violate this condition.

##### B.2 Schedule of Moves

The schedule of moves depends on how frequently we wish to execute local moves as opposed to global ones. We capture this ratio using the fixed, user-specifed parameter $M_{\text{local}}$, which states that there are $M_{\text{local}}$ iterations of each local move per 1 iteration of each global move. Letting $M_{\text{global}}$ denote the total number of iterations of each global move and $M_{\text{record}}$ be the number of local moves per MCMC sample (i.e. state of the MCMC that is returned to the user at the end of the algorithm), we propose Algorithm 1 as the overall structure of our implementation. As a default, we propose $M_{\text{global}} = 10^4$, $M_{\text{local}} = 100$, and $M_{\text{record}} = 100$.

**Algorithm 1** Phylogenetic and Transmission Reconstruction

Initialize $\boldsymbol{\theta}$, the configuration of the transmission network and values of all parameters
$\boldsymbol{\Theta} \leftarrow \emptyset$, the set of posterior samples
**for** $1 \leq i \leq M_{\text{global}}$ **do**
    Update $\boldsymbol{\theta}$ by executing moves 1-3 in order
    **for** $1 \leq j \leq M_{\text{local}}$ **do**
        Update $\boldsymbol{\theta}$ by executing moves 4-16 in order
        **if** $j \bmod M_{\text{record}} = 0$ **then**
            Append $\boldsymbol{\theta}$ to $\boldsymbol{\Theta}$
        **end if**
    **end for**
**end for**
**return** $\boldsymbol{\Theta}$

##### B.3 Parallelization

The above MCMC sampler may conveniently be run in parallel over subtrees partitioning the transmission network, thanks to the fact that the likelihood function computed on the entire tree equals the product of the likelihood function computed on each subtree. The only moves that require an update of the likelihood on all parts of the tree at once are the global moves; hence, we parallelize the algorithm by randomly partitioning the tree into subtrees after completing the global moves, then joining the subtrees back together to perform the next cycle of global moves, and repeating.[1]

[1] Moves 10 and 11 do sometimes change the likelihood across different regions of the transmission network; to resolve this, we simply limit the choices of the new value of $h_i$ to those within the same subtree as $i$. Moreover, we

To perform the random tree partitioning, we implement the algorithm presented in Borndörfer, Elijazyfer, & Schwartz (2019), with a slight modification. In words, this algorithm first specifies $\lambda$, the minimum allowable number of nodes in a subtree. Here we use a conservative choice of $\lambda = \max(n/M_{\text{cores}}, 25)$, where $n$ is the total number of nodes in the original tree and $M_{\text{cores}}$ is the available number of CPU cores. This choice guarantees that the algorithm will produce at most $M_{\text{cores}}$ subtrees. Then, we iterate over nodes in the tree in reverse-BFS order, where BFS is executed starting at the root node. For a node $i$, we first compute the total number of descendants of $i$ as the sum of the number of descendants of the children of $i$, plus 1 for $i$ itself. After having performed this update, if the total number of descendants of a node $i$ is $\lambda$ or greater, we take $i$ to be the root of one of our subtrees—so long as "cutting off" that subtree leaves at least $\lambda$ nodes remaining in the subtree rooted at the global root. Finally, for the sake of improved mixing, we make the modification that a node $i$ cannot be the root of a subtree if it was the root of a subtree in the previous partition of the transmission network.

automatically reject any move that affects a host $i$ at the boundary of one subtree and another and would hence cause the likelihood to change in more than one subtree. Since each tree partition differs from the previous one and varies randomly with the tree topology, a certain tree rearrangement via Move 10 or 11 prohibited under one partition may, and eventually will, become possible under a different partition.
